# Clinical Validation of RlapsRisk BC in an international multi-cohorts setting

**DOI:** 10.1101/2025.07.18.25331788

**Authors:** V. Gaury, Z. Vaquette, V. Aubert, C. Barcenas, L.J. Jones, D. Jacobs, J. Guillon, A. Filiot, B. van der Vegt, E. Hocquet, C. Maisin, L.K. Spary, M. Bateson, A.-L. Martin, D. Drubay, S. Everhard, L. Guillou, M. Sefta, I. Garberis, F. André, A. Vincent Salomon, S. Krishnamurthy, M. Lacroix-Triki

## Abstract

**Purpose:** This study evaluated the prognostic performance of RlapsRisk BC, a multimodal deep learning tool designed to predict distant recurrence-free interval (dRFI) in early-stage, ER-positive, HER2-negative breast cancer using routine H&E-stained whole-slide images (WSIs) and standard clinicopathologic features.

**Methods:** RlapsRisk BC was developed and internally validated on seven retrospective cohorts totaling 6,039 patients. Its ability to stratify dRFI risk was validated retrospectively in 594 ER+/HER2− breast cancer patients across three international cohorts.

**Results:** RlapsRisk BC effectively stratified patients into low- and high-risk groups (HR 3.93–9.02), with 5-year distant recurrence of 0.85%–4.73% vs. 6.26%–34.74%, consistently across key subgroups. RlapsRisk BC was an independent prognostic factor, improving the c-index by 0.05 to 0.19 when combined with clinical variables, with significant gains in two cohorts.

Compared to genomic assays, RlapsRisk BC showed complementary—and sometimes superior—performance. At matched specificity, it achieved higher sensitivity: 0.85 vs. 0.33 (Oncotype DX) and 0.74 vs. 0.49 (EndoPredict).

**Conclusion:** RlapsRisk BC demonstrates strong, independent prognostic value and may offer a scalable, accessible alternative to genomic assays. Further studies are needed to confirm clinical utility and support integration into treatment decisions.

## Background

Estrogen receptor–positive (ER+), human epidermal growth factor receptor 2–negative (HER2−) breast cancer accounts for approximately 70% of early-stage breast cancer (eBC) cases and encompasses a heterogeneous group of tumors with varying risks of recurrence. While most patients benefit from endocrine therapy and have favorable short-term outcomes, particularly those with luminal A–like tumors, a substantial subset remains at risk for late distant relapse, often occurring more than a decade after initial treatment [1]. Accurately identifying patients at high or low risk of recurrence is critical to optimizing adjuvant treatment, avoiding unnecessary chemotherapy, and considering extended or intensified endocrine strategies when appropriate [2].

Risk stratification is currently guided by clinicopathologic features, integrated into tools such as the Nottingham Prognostic Index and PredictBreast [3], and increasingly supported by molecular assays like Oncotype DX, MammaPrint, EndoPredict, and Prosigna. While these multigene assays improve prognostic precision, they are associated with practical limitations, including cost, accessibility, and turnaround time, that may restrict their widespread use. Consequently, there is a need for scalable alternatives that maintain or exceed the performance of existing methods.

In a previous proof-of-concept study, we introduced RlapsRisk BC, a multimodal deep learning–based prognostic tool that integrates features extracted from hematoxylin and eosin (H&E)–stained whole-slide images (WSIs) with routine clinical data to estimate the risk of distant recurrence in ER+/HER2− eBC [4]. Initial findings demonstrated that image-based models can capture global biologically meaningful prognostic information ascertained from the entire tissue section and offer a viable path toward accessible, inherent pathology-based biomarkers.

In the present study, we expand upon that work by conducting a large-scale, blinded, retrospective validation of RlapsRisk BC in three independent, international cohorts comprising patients treated with endocrine therapy alone, a clinically important subgroup for considering the potential addition of chemotherapy. Currently, several molecular tests, such as Oncotype DX or EndoPredict, estimate the risk of distant recurrence, but the groups of high- and low-risk patients vary from one test to another. Here, in addition to assessing the model’s standalone prognostic performance, we directly compare RlapsRisk BC with some of the commercially available genomic assays, to evaluate whether this AI-based approach can outperform or complement them in identifying patients who may safely avoid chemotherapy (Figure 1).

**Figure 1:**
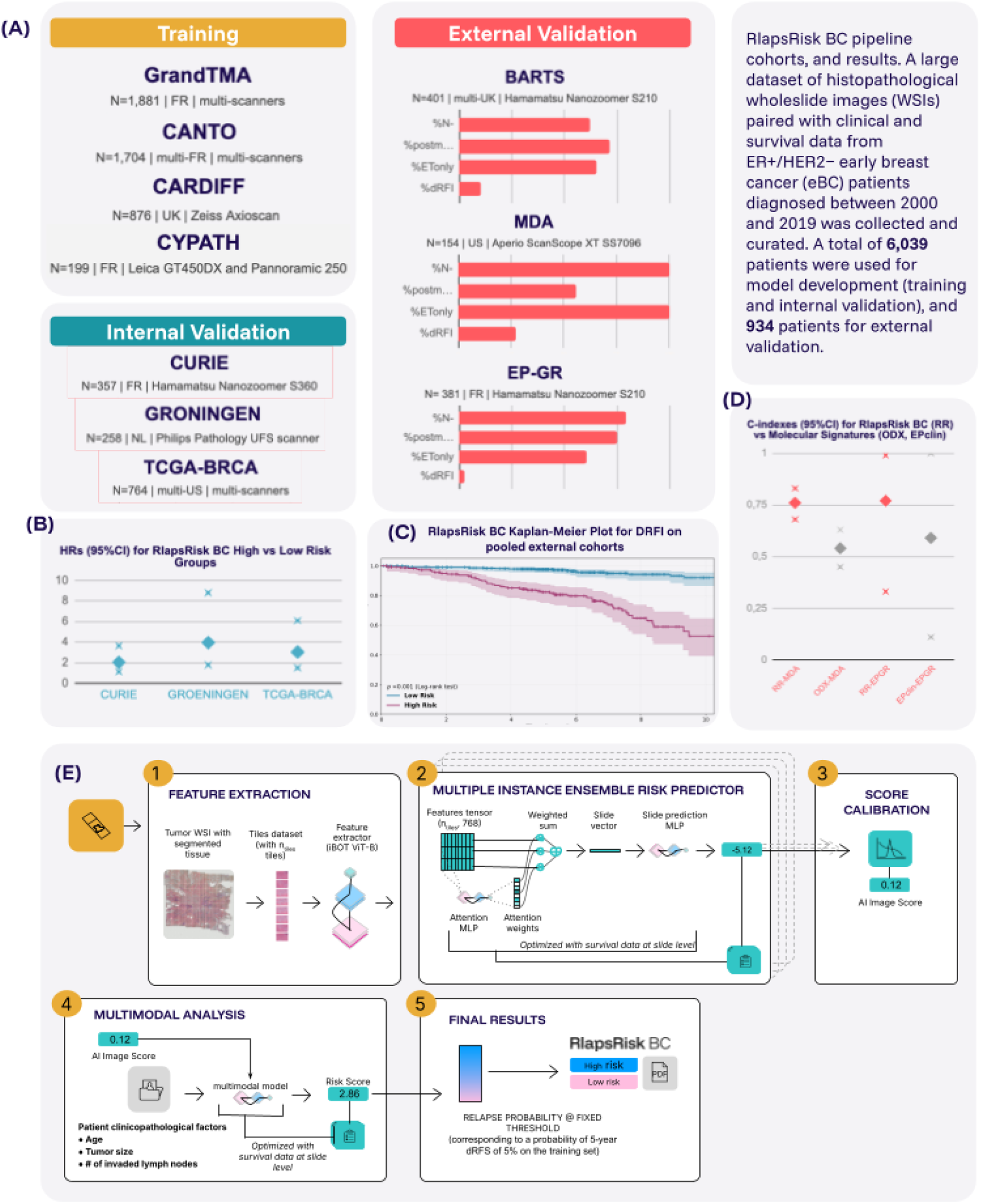
Schematic overview of the RlapsRisk BC pipeline cohorts, and results. A large dataset of histopathological whole-slide images (WSIs) paired with clinical and survival data from HR+/HER2− early breast cancer (eBC) patients diagnosed between 2000 and 2019 was collected and curated. A total of 6,039 patients were used for model development (training and internal validation), and 934 patients for external validation. **(A)** Cohorts overview: training (blue), internal validation (teal), and external validation (pink), with sample sizes, countries, scanner models, and key clinical characteristics (percentages of: node-negative patients, post-menopausal patients, patients treated with endocrine therapy only, dRFI events). **(B)** Hazard ratios (HRs) comparing RlapsRisk BC high- vs. low-risk groups in internal validation sets. **(C)** Kaplan–Meier survival curve showing RlapsRisk BC stratification for dRFI on pooled external validation cohorts for patients treated with endocrine therapy only (N=594). **(D)** C-indices of RlapsRisk BC versus existing molecular scores for patients treated with endocrine therapy only on MDA (Oncotype Dx, N=154) and EP-GR (EPclin, N=180). **(E)** RlapsRisk BC’s pipeline scheme: WSI-based feature extraction, multiple instance learning risk prediction, score calibration, integration with clinical variables, and final risk stratification. Abbreviations: C-index, concordance index; dRFI, distant recurrence-free interval; HR⁺/HER2⁻, hormone receptor–positive, HER2-negative; WSI, Whole Slide Image.

**Table 1:**
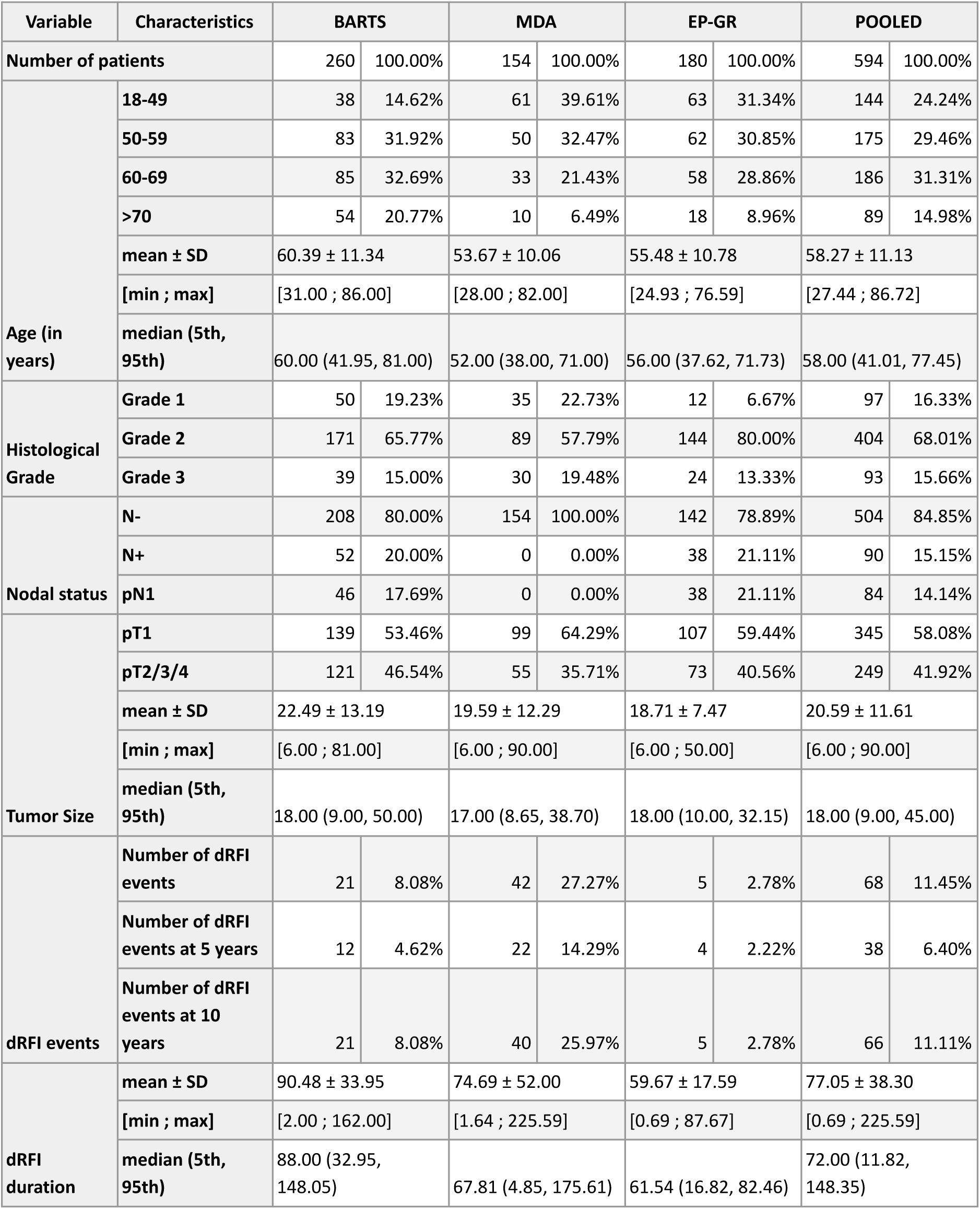
Characteristics of the three cohorts used to evaluate RlapsRisk BC performances, for the population treated with adjuvant endocrine therapy only. Detailed description of the cohorts stratified by treatment regimen can be found in the supplementary material. Abbreviations: dRFI: distant recurrence-free interval.

## Methods

### Study Design & Setting

This study reports the blind external evaluation of RlapsRisk BC, a multimodal deep learning–based prognostic model developed to estimate the risk of distant recurrence in ER+/HER2− early breast cancer. The model was trained to directly predict distant recurrence-free interval (dRFI) using retrospective multicenter data combining digitized hematoxylin and eosin (H&E) stained slides with routinely collected clinicopathologic variables (age, tumor size, and number of invaded lymph nodes).

We performed a blinded evaluation of RlapsRisk BC across three independent retrospective cohorts: BARTS (UK), MDA (USA), and EP-GR (France). These cohorts were not involved in the model training and were selected to ensure geographic, institutional, and population diversity.

The primary endpoint was dRFI at five years. Secondary analysis included stratification by clinical subgroups and comparison with existing clinical and genomic prognostic tools, where available.

### Endpoint Definition

dRFI is defined as the time from definitive surgery to the first occurrence of distant metastatic recurrence, with patients censored at the time of death from other causes or last follow-up if no distant event has occurred. This endpoint specifically captures the development of metastatic disease and excludes locoregional recurrence or contralateral breast cancer, thereby providing a focused measure of distant disease control.

### AI Model Development

A first version of the RlapsRisk BC was designed to predict 5-year distant relapse directly from hematoxylin, eosin, and saffron (HES) stained WSIs, without requiring pathologist annotations [4]. In the present study, a new architecture and training procedures are used, where optimized losses were introduced and a more performant and robust feature extractor implemented. The neural network produces a digital pathology score from WSIs, which is then combined with three clinicopathological variables – age, number of invaded lymph nodes, and tumor size – using a Weibull survival model to generate RlapsRisk BC score, a prognostic indicator of dRFI.

A large dataset of histopathological whole-slide images (WSIs) paired with clinical and survival data from ER+/HER2− eBC patients diagnosed between 2000 and 2019 was collected, curated and leveraged to develop this new version. A total of 6,039 patients were included for model development (training and internal evaluation) (**Figure 1**). The goal was to train a multimodal deep learning model to predict dRFI directly from WSIs and standard prognostic clinical variables. Model training was performed using data from the GrandTMA [5] (n=1,881), CANTO [6] (n=1,704), CYPATH (n=199), and CARDIFF (n=876) cohorts.

Internal performance assessment was conducted using three retrospective cohorts: TCGA-BRCA (n = 764), GRONINGEN (n = 258), and CURIE (n = 357). Prognostic performance was evaluated through stratified bootstrap evaluation (N=5,000). Full details on cohort characteristics, model architecture, and training procedures are provided in the Supplementary Materials.

To ensure that the model was able to produce a meaningful prediction, a minimum amount of tumor tissue was required to run the algorithm: after dividing the WSI in 112μm x 112μm image patches (tiles), only slides with at least 150 tiles (approx. 1.9 mm^2^) containing tumor were kept. As a result, 25 slides from TCGA and 4 slides from BARTS were excluded from the analyses.

### Study population

All patients underwent surgical resection as initial treatment and had confirmed ER+/HER2− early breast cancer. Inclusion required follow-up data and at least one digitized H&E–stained WSI of the primary tumor (diagnostic slide when available). Clinical variables for risk computation—age, tumor size, and number of positive lymph nodes—had to be available (inclusion/exclusion flowchart in **Supplementary Figure 1)**.

Clinical validation was performed on three independent external cohorts: MDA (n=154), BARTS (n=399), and EP-GR (n=381). Model performance was assessed per cohort and in the combined POOLED dataset (n=934).

Molecular signature results were available for three cohorts: EndoPredict (EPclin) in EP-GR; Oncotype DX (RS) in MDA and OD-GR (n=161), the latter lacking follow-up data. Evaluation focused primarily on patients treated with endocrine therapy alone (full cohort characteristics in Supplementary Materials).

### Statistical Analyses

The primary endpoint was dRFI, defined per STEEP criteria [7] as the time from breast cancer diagnosis to the first distant recurrence or death from breast cancer. Outcomes such as local–regional recurrence, ipsilateral or contralateral ductal carcinoma in situ (DCIS), invasive contralateral breast cancer, and second non-breast primaries were not counted as events.

To evaluate prognostic performance, we used the IPCW C-index and AUC from scikit-survival (v0.19.0), accounting for right-censoring. AUC comparisons used the Blanche et al. method via the timeROC R package. Patients were stratified using a 5% 5-year relapse probability threshold, derived from a 10% 10-year dRFI exponential model, consistent with clinical molecular signatures. Kaplan–Meier and log-rank tests compared survival between risk groups. Cox models assessed the model’s independent prognostic value. Sensitivity, specificity, PPV, and NPV (via timeROC) were calculated for 5-year distant recurrence. Wald chi-square tests and hazard ratios quantified variable impacts. The methods to describe the Other prognostic methods used for comparison are explained in the supplementary material.

## Results

### Internal Evaluation

In the internal validation, RlapsRisk BC demonstrated consistent prognostic performance across the three internal cohorts. The concordance indices were 0.70 for TCGA-BRCA, 0.69 for CURIE, and 0.69 for GRONINGEN, indicating good discriminative ability. Stratification into high- and low-risk groups yielded hazard ratios of 3.04 (95% CI, 1.52–6.09; p = 0.002) in TCGA-BRCA, 2.03 (95% CI, 1.13–3.64; p = 0.018) in CURIE, and 3.96 (95% CI, 1.79–8.78; p < 0.001) in GRONINGEN, supporting the reproducibility of RlapsRisk BC’s risk predictions across independent internal datasets after models’ parameters selection.

### Cohort characteristics

To evaluate the prognostic performance of the RlapsRisk BC model, we performed an external validation using three independent retrospective cohorts with available outcome data, including a total of 934 patients with ER+/HER2– eBC: MDA (n = 154), BARTS (n = 399), and EP-GR (n = 381). Among them, 594 patients received adjuvant endocrine therapy alone. A fourth cohort, OGDR (n = 267), for which no follow-up data were available, was included separately only for further concordance assessment between RlapsRisk BC and Oncotype DX. The combined dataset of the three cohorts with survival data is referred to as the POOLED cohort. Molecular signature test results were available for EndoPredict in the EP-GR cohort, and for Oncotype DX in both the MDA and OD-GR cohorts. Summary statistics for full cohorts, including patients who received chemotherapy, are provided in Supplementary Material (**Table S1-S2**). The cohorts’ characteristics presented in this section focus exclusively on patients treated with five years of adjuvant endocrine therapy alone (see supplementary for characteristics of chemotherapy-treated patients).

In the BARTS cohort (n = 399), 260 patients (65.2%) received adjuvant endocrine therapy without chemotherapy. The mean age in this subgroup was 60.4 years (SD 11.3), and 80.4% of patients were postmenopausal. The mean tumor size was 22.5 mm (SD 13.2). 80% of cases had no lymph node involvement. Histological grade was predominantly low to intermediate: 19.2% of tumors were grade 1 and 65.8% were grade 2. During a median follow-up of 88 months (SD 34.0), 21 dRFI events occurred in this subgroup (recurrence rate: 8%).

The MDA cohort included 154 node-negative patients, all treated with adjuvant endocrine therapy alone. It was specifically enriched with patients who experienced distant metastasis within 5 years, each matched to a patient who remained metastasis-free during the same period. The mean age was 53.7 years (SD 10.1), and 55.8% of patients were premenopausal. The average tumor size was 19.6 mm (SD 12.3). Histological grading showed 22.7% grade 1 and 52.8% grade 2 tumors. Over a median follow-up of 67.8 months (SD 52.0), 42 dRFI events were recorded (27.3%).

In the EP-GR cohort (n = 381), 180 patients (47.2%) were treated with endocrine therapy alone. Their mean age was 59.1 years (SD 10.6), and 72.2% were postmenopausal. The average tumor size was 18.7 mm (SD 7.5). 78.9% of patients were node-negative. Histological grades were distributed as follows: 6.7% grade 1, 80% grade 2, and 13.3% grade 3. The mean follow-up duration was 59.4 months (SD 17.6), and 5 dRFI events were observed (recurrence rate: 2.8%).

The OD-GR cohort (n = 267), for which follow-up data were not available, was used exclusively for a concordance analysis with Oncotype DX. Among these patients, 161 (60.2%) received endocrine therapy alone. Their mean age was 58.8 years (SD 7.8), and 81.4% were postmenopausal. The mean tumor size in this subgroup was 21.6 mm (SD 10.5). Node-negative disease was present in 36.6% of patients. Tumor grade was predominantly intermediate (77.6% grade 2, 13.0% grade 3).

All main results refer to patients treated with five years of adjuvant endocrine therapy alone and with adequate follow-up, except the final Results paragraph, which includes those who also received chemotherapy. Focusing on endocrine therapy alone allows a clearer evaluation of the score’s prognostic value in identifying low-risk patients who might safely avoid chemotherapy. For completeness, we also present findings from the combined external validation cohorts (the POOLED dataset).

### Prognosis performance

Across the three independent external validation cohorts, RlapsRisk BC demonstrated a significant stratification of dRFI between low- and high-risk subgroups (P < .0005; **Figure 2**). The model achieved a concordance index (c-index) of 0.71 (95% CI, 0.59–0.82) with a hazard ratio (HR) of 3.93 (95% CI, 1.65–9.34; p=0.002) in the BARTS cohort; a c-index of 0.76 (95% CI, 0.68–0.83) and HR of 7.54 (95% CI, 3.34-16.99; p=<0.001) in the MDA cohort; and a c-index of 0.77 (95% CI, 0.33-0.99) and HR of 9.02 (95% CI, 0.99-82.09; p=0.051) in the EP-GR cohort. In the pooled analysis, performance reached a c-index of 0.77 (95% CI, 0.71-0.84) and HR of 7.44 (95% CI, 4.33-12.78; p=<0.001).

**Figure 2:**
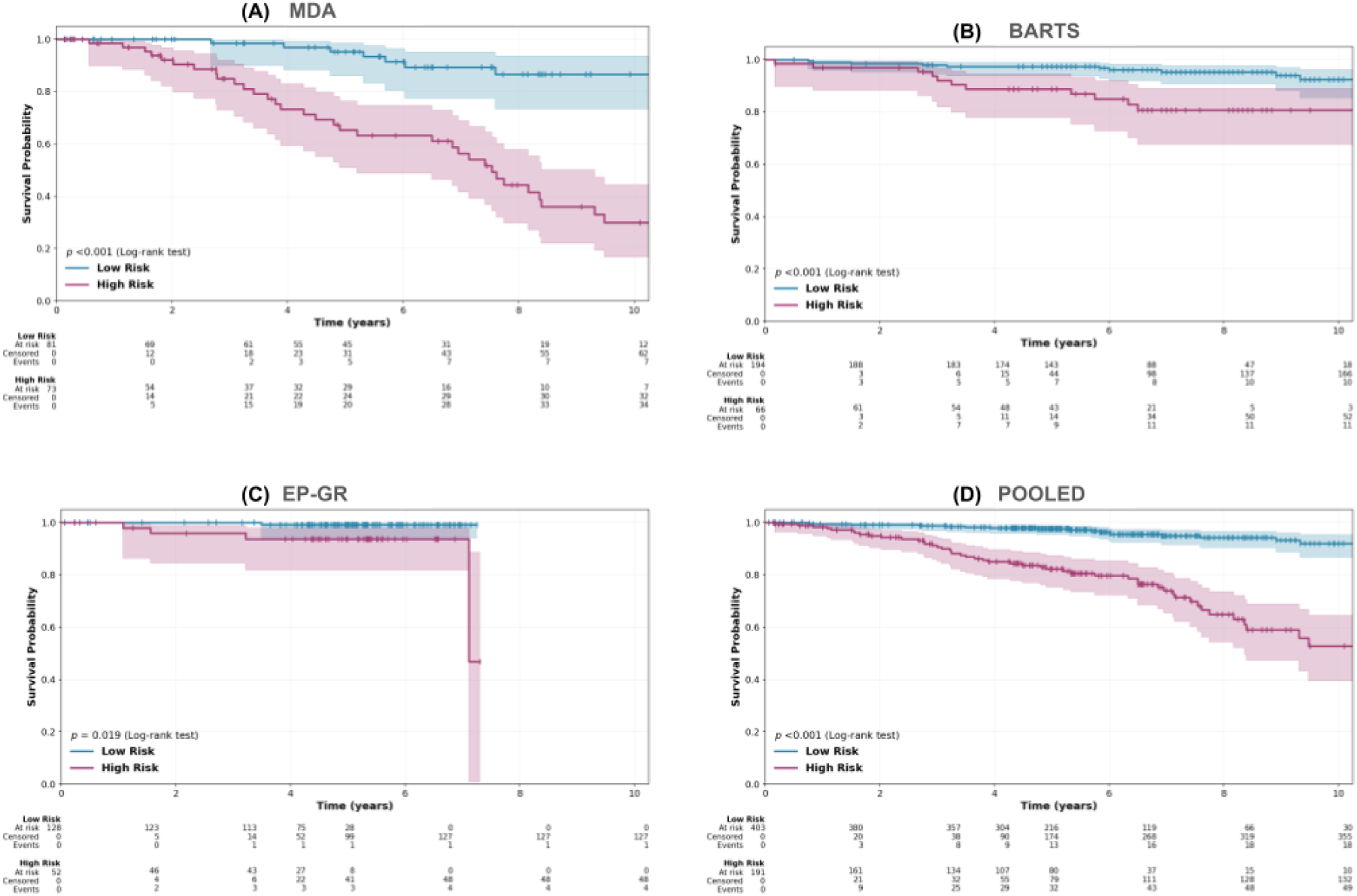
Kaplan-Meier curves for the dRFI endpoint in HR+/HER2- eBC treated with endocrine therapy only, for the high and low-risk subgroups provided by RlapsRisk BC. **(A)** The upper left panel corresponds to the MDA cohort, **(B)** the upper right panel corresponds to the BARTS cohort, **(C)** the lower left panel corresponds to the EP-GR cohort, and **(D)** the lower right panel corresponds to the POOLED cohort. Stratification p-values were computed with a logrank test.

At five years, the predicted low-risk subgroups exhibited low probabilities of dRFI events, always below a value of 5%: 2.61% (95% CI, 1.0%–6.2%) for the BARTS cohort, 4.73% (95% CI, 1.55%–13.98%) for the MDA cohort, 0.85% (95% CI, 0.12%–5,86%) for the EP-GR cohort, and 2.41% (95% CI, 1.26%–4.58%) in the pooled analysis. In contrast, the high-risk subgroups showed substantially higher 5-year dRFI event probabilities: 11.24% (95% CI, 5.52%–22.16%) for BARTS, 34.74% (95% CI, 23.63%–49.12%) for MDA, 6.26% (95% CI, 2.06%–18.16%) for EP-GR, and 17.80% (95% CI, 12.70%–24.65%) in the POOLED cohort.

When restricting the analysis to multiple clinically relevant subgroups, the difference in terms of dRFI events between the high- and low-risk groups remained statistically significant (P ≤ 0.005, **Figure 3** and **Supplementary Figure 2**), notably for histological grade 2 patients, pre-and post-menopausal status, as well as N0 patients (dRFI event rates respectively for BARTS, MDA, EP-GR, POOLED in % for Grade 2: 7.6%, 29.2%, 3.5%, 10.6%; pre-menopausal: 11.8%, 22.1%, 2.0%, 13.0%; post-menopausal: 7.2%, 31.4%, 3.1%, 10.8%; N0: 6.3%, 27.2%, 2.8%, 11.7%). The model demonstrated consistent prognostic value across these subgroups, including those in which risk estimation is typically more uncertain. For instance, node-negative and small tumors (pT1) are usually considered lower risk, yet the model was able to identify patients at substantially higher risk within these groups, suggesting a role in refining adjuvant treatment decisions. Similarly, robust discrimination was observed in the intermediate Oncotype DX group, where conventional genomic testing often leaves clinical ambiguity, highlighting the model’s potential to complement existing tools. In the POOLED cohort, most HRs across subgroups were significant, (**Figure 3**, left panel) with values ranging from 1.28 (95% CI, 1.07 to 1.54) to higher values. Notable exceptions are grade 1 patients, and invasive lobular cancers where the sample size and the number of events were limited, resulting in very large confidence intervals. Apart from invasive lobular cancers (0.64, 95% CI, 0.14 to 0.95), the concordance index (c-index) across subgroups ranged from 0.72 (95% CI, 0.67 to 0.86) to 0.82 (95% CI, 0.74 to 0.90), supporting the model’s robust discriminatory performance across diverse clinical contexts (**Figure 3**, right panel).

**Figure 3:**
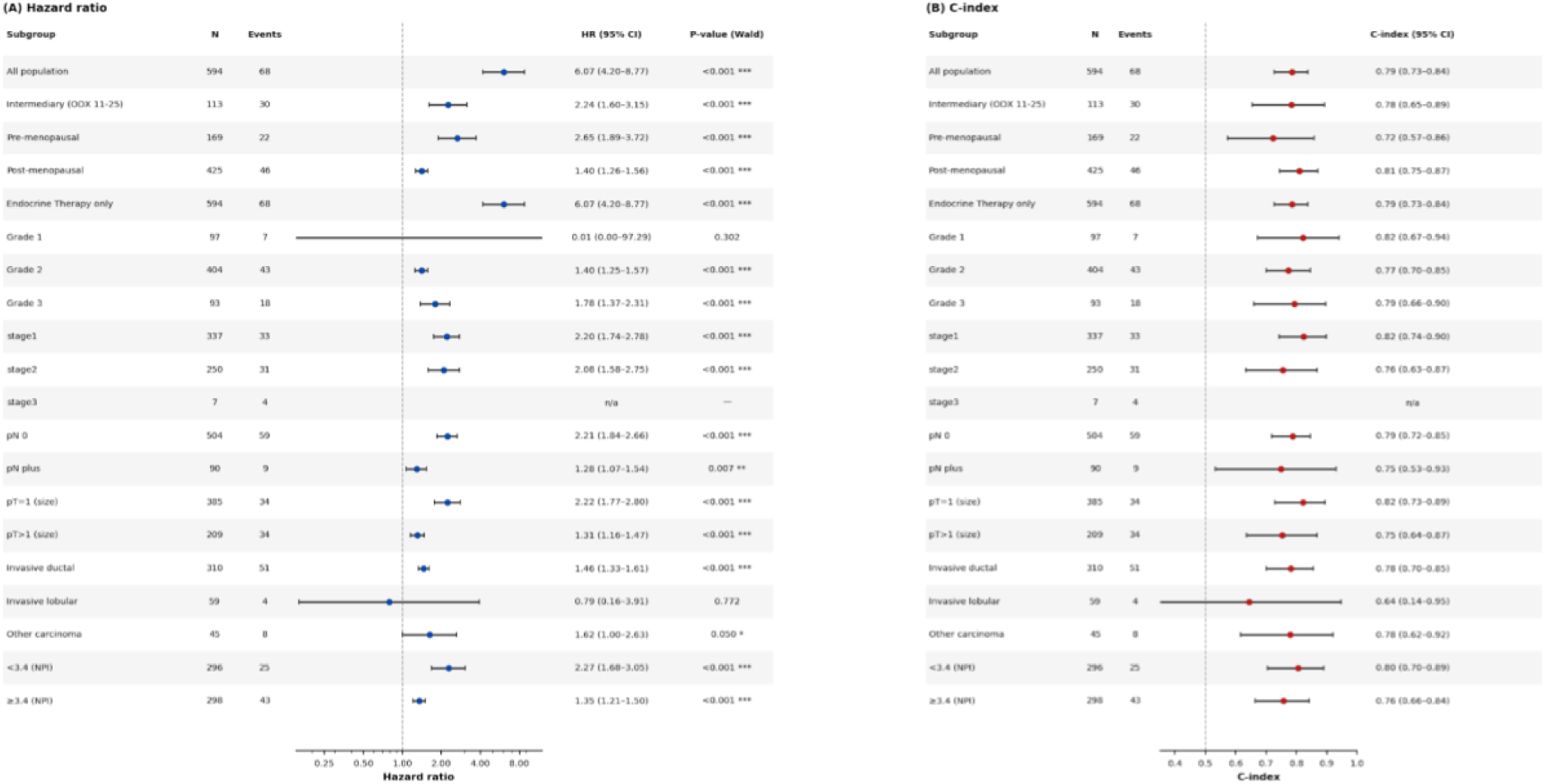
Prognostic performance of RlapsRisk BC across clinical subgroups in the pooled external validation cohort. This figure presents two panels evaluating the prognostic value of the RlapsRisk BC score in predefined subgroups of patients with hormone receptor–positive, HER2-negative (HR⁺/HER2⁻) breast cancer. **(A)** The left panel shows a forest plot of the hazard ratio (HR) for distant recurrence-free interval (dRFI) comparing high- versus low-risk groups, also with 95% CIs, derived from univariate Cox proportional hazards models. **(B)** The right panel displays a forest plot of the concordance index (C-index) with corresponding 95% confidence intervals (CIs), indicating the discriminative accuracy of the score within each subgroup. Subgroups analyzed include Oncotype DX risk groups (high vs low), menopausal status, treatment (endocrine therapy alone vs endocrine therapy and chemotherapy), histologic grade (I–II vs. III), tumor stage, nodal status (N0 vs. N+), tumor size (pT1 vs. pT2+), histologic subtype, and NPI (above or below 3.4). RlapsRisk BC demonstrated consistent prognostic value across these clinically relevant strata, with high C-index and significantly elevated HRs for high-risk patients across most subgroups. HR p-values were computed with a Wald test. Abbreviations: HR, hazard ratio; CI, confidence interval; C-index, concordance index; dRFI, distant recurrence-free interval; HR⁺/HER2⁻, hormone receptor–positive, HER2-negative.

### Comparison with molecular and clinical prognostic scores

#### Clinical factors

We found that the AI-derived score generated from Whole Slide Images (WSIs) provides independent prognostic information beyond established clinical variables, including histological grade, patient age, lymph node involvement, tumor size, and Ki-67 expression – although Ki-67 data were not available in the BARTS cohorts, missing values were replaced with the median values computed from the other cohorts (P ≤ 0.005). Integrating RlapsRisk BC with clinical variables (tumor size, nodes, age) improved 5-year dRFI prediction. In the MDA cohort, the combined model yielded a c-index of 0.76 (95% CI: 0.68–0.83) vs. 0.57 (95% CI: 0.48–0.66) for clinical factors alone (P = 0.013; **Supplementary Table 5**). Similarly, in BARTS and in EP-GR, RlapsRisk BC outperformed clinical factors (0.71 [95% CI: 0.59–0.82] vs. 0.68 [95% CI: 0.57–0.79] and 0.77 [95% CI: 0.33–0.99] vs. 0.72 [95% CI: 0.46–0.97]). In the POOLED cohort, RlapsRisk BC achieved a c-index of 0.77 (95% CI: 0.71–0.84), compared to 0.58 (95% CI: 0.51–0.65) for clinical variables alone.

In the POOLED cohort, RlapsRisk BC outperformed established prognostic models, achieving a c-index of 0.77 (95% CI: 0.71–0.84) versus 0.60 (95% CI: 0.53-0.68) for CTS5 and 0.52 ((95% CI: 0.43-0.60) for PredictBreast. It also outperformed clinical factors across key subgroups (grade, menopausal status, nodal involvement; see **Supplementary Table 3**). Compared to CTS5, RlapsRisk BC showed better classification metrics: PPV of 0.182 vs. 0.156, NPV of 0.976 vs. 0.936, and more balanced sensitivity and specificity (both 0.74 vs. 0.17 and 0.92, respectively).

#### Molecular Signatures

We next compared the Oncotype DX Recurrence Score (RS) and the RlapsRisk BC score within the MDA cohort, where RS data were available. The two scores showed no significant correlation (r = 0.15, p = 0.072), and concordance in risk classification was modest at 55.20%, with high-risk RS patients defined using the thresholds defined in the Methods section (**Supplementary Figure 4**, upper panel). Similarly, in the OD-GR cohort, the scores were not correlated (r = 0.15, p = 0.086), with a concordance of 73%, higher than from the MDA cohort (**Supplementary Figure 4**, lower panel).

We compared the prognostic performance of RlapsRisk BC with that of the Oncotype DX (RS) classification for predicting 5-year dRFI in the MDA cohort, using the thresholds defined in the Methods section. C-index for continuous scores was significantly superior for RlapsRisk BC (0.76, 95% CI: 0.68–0.83) compared to RS (0.54, 95% CI: 0.45–0.63).

RlapsRisk BC provided a more pronounced separation between high- and low-risk patients (HR = 7.54; 95% CI, 3.34–16.99, p=<0.001) compared to RS (HR = 0.98; 95% CI, 0.53-1.83, p=0.959) (**Figure 4** upper panel). RlapsRisk BC achieved a sensitivity of 0.86 and a specificity of 0.63, while RS reached a sensitivity of 0.33 and a specificity of 0.61. Positive predictive value (PPV) was at 0.34 for RlapsRisk and 0.16 for RS, yet both achieved high negative predictive values (NPV) of 0.95 and 0.80, respectively (**Supplementary Table 4**).

**Figure 4:**
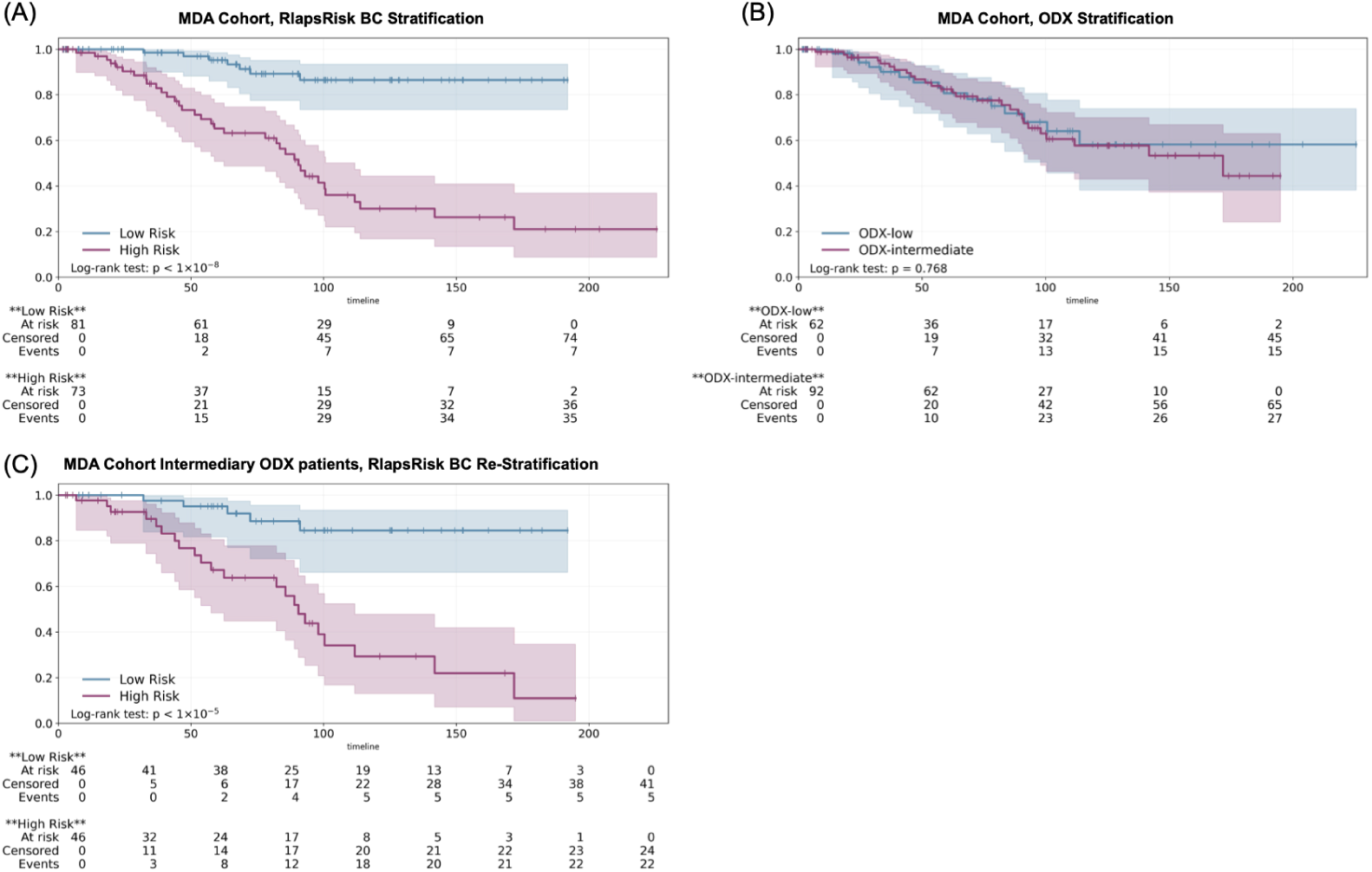
Kaplan-Meier curves for the dRFI endpoint in HR⁺/HER2⁻ eBC treated with endocrine therapy only in the MDA cohort. **(A)** The upper left panel displays risk group stratification using RlapsRisk BC, while **(B)** the upper right panel shows stratification based on the Oncotype DX Recurrence Score (RS). **(C)** The lower panel illustrates re-stratification of patients classified as intermediate-risk by RS using the RlapsRisk BC score. Stratification p-values were computed with a logrank test. Abbreviations: dRFI, distant recurrence-free interval; HR⁺/HER2⁻, hormone receptor–positive, HER2-negative.

To assess the potential of RlapsRisk BC to refine risk assessment in patients classified as intermediate risk by Oncotype DX, we conducted a focused analysis within this subgroup. RlapsRisk BC further stratified these patients into two groups with significantly different outcomes (**Figure 4** lower panel, HR=10.5 (3.1–34.2), p< 0.005).

In the EP-GR cohort, we compared RlapsRisk BC to EPclin scores in 180 patients treated with endocrine therapy only where both scores were available. The two scores were significantly correlated with a correlation coefficient r of 0.33 (p<0.001). The proportion of patients classified as high-risk was similar between models: RlapsRisk BC identified 28.9% of patients as high-risk, compared to 31.7% for EPclin (**Supplementary Figure 5**). Yet RlapsRisk BC demonstrated superior discriminative performance with a c-index of 0.77 (95% CI, 0.33–0.99) versus 0.59 (95% CI, 0.11–1) for EPclin.

Additionally, the two models differed in their classification performance. RlapsRisk BC achieved a sensitivity of 0.74 and a specificity of 0.74, while EPclin reached a sensitivity of 0.49 and a specificity of 0.72. Positive predictive value (PPV) was modest for both scores, at 0.063 for RlapsRisk and 0.057 for EPclin, yet both achieved high negative predictive values (NPV) of 0.991 and 0.984, respectively (**Supplementary Table 4**).

### Performance in Chemotherapy-Treated Patients

To further evaluate the robustness of RlapsRisk BC in a higher-risk clinical setting, we assessed its prognostic and classification performance in patients who received both endocrine therapy and chemotherapy within the POOLED cohort. In this population, RlapsRisk BC stratified patients into two distinct risk groups, but statistical significance was not reached in distant recurrence risk (HR = 2.13; p =0.078; **Supplementary Figure 6**). At 5 years, the incidence of distant recurrence was 2.7% (95% CI, 1.1%–6.3%) in the low-risk group compared to 7.1% (95% CI, 3.7%–13.2%) in the high-risk group. The overall prognostic performance of RlapsRisk BC was comparable to that of established clinical scores, with a C-index of 0.71 (95% CI, 0.57–0.84), compared to 0.73 (95% CI, 0.62–0.83) for CTS5 and 0.65 (95% CI, 0.56–0.74) for PredictBreast.

In terms of classification performance, RlapsRisk BC achieved a sensitivity of 0.64 (± 0.12) and a specificity of 0.60 (± 0.03), while CTS5 showed a sensitivity of 0.56 (± 0.13) and a specificity of 0.69 (± 0.03). Positive predictive values were similarly low for both scores, with 0.071 (± 0.02) for RlapsRisk BC and 0.081 (± 0.02) for CTS5. Negative predictive values were high and nearly identical, at 0.973 (± 0.01) for RlapsRisk BC and 0.971 (± 0.01) for CTS5.

## Discussion

This study presents a blinded validation of RlapsRisk BC, an AI-based prognostic model that integrates whole-slide histopathological features with standard clinicopathologic variables (age, tumor size, and positive lymph nodes). Validation was conducted in three independent retrospective cohorts of ER-positive, HER2-negative early breast cancer, along with a pooled analysis. Main results focus on patients treated with 5 years of adjuvant endocrine therapy alone—the standard baseline treatment—providing a consistent, clinically relevant context to assess the model’s prognostic value.

Across all cohorts, RlapsRisk BC consistently demonstrated significant prognostic performance for dRFI. The model stratified patients into clearly distinct low- and high-risk groups, with hazard ratios (HRs) ranging from 3.93 to 9.02 across validation cohorts. At 5 years, the probability of distant recurrence in low-risk patients ranged from 0.85% to 4.73%, compared to 6.26% to 34.7% in high-risk patients. The separation of risk groups was consistent across key clinical subgroups, including histological grade 2 and pre-menopausal patients.

RlapsRisk BC integrates standard clinical variables that are robust and routinely collected, ensuring consistent applicability across healthcare settings. By combining these with digitized routinely available H&E slides, the model provides prognostic information that is complementary to traditional clinico-pathologic features. In all cohorts, RlapsRisk BC achieved a superior c-index compared to the score based on clinical variables alone.

Comparative analyses with genomic assays further demonstrate the potential utility of RlapsRisk BC. It showed no correlation with the Oncotype DX recurrence score, indicating that it captures distinct prognostic signals. RlapsRisk BC provided additional granularity by reclassifying patients with intermediate Oncotype DX recurrence scores into groups with distinct outcomes. Compared to EndoPredict, it identified fewer patients as high-risk while maintaining a higher negative predictive value, suggesting improved ability to reassure patients with truly low risk of distant recurrence. Overall, this ability to improve risk stratification makes it a promising tissue-efficient alternative or complementary option to current genomic assays such as Oncotype DX and EndoPredict in ER+/HER2– early breast cancer.

While initial comparisons are encouraging, they should be interpreted cautiously due to potential confounding, particularly from treatment decisions influenced by genomic assay results. Further studies comparing RlapsRisk BC to molecular signatures are needed to clarify its role in clinical decision-making and assess its use as a surrogate or complement to existing assays. Recent work supports the potential of histology-based AI models in ER+/HER2− early breast cancer: Lee et al. showed that features from H&E slides can predict high genomic risk with strong validation on TCGA data [14]; the Ataraxis test, trained on over 8,000 patients, outperformed genomic assays in HR+ cohorts (C-index 0.67 vs. 0.61) [15]; and the Orpheus model achieved an AUC of 0.89 for identifying patients with recurrence scores >25 [16]. These studies underscore the growing promise of AI-driven pathology as a viable alternative to molecular assays. Another limitation of the present work is the relatively small size of the validation cohorts, in particular when it comes to subgroup analyses. Although several of the results presented here are significant, further explorations with larger cohorts are required to conclude on the prognostic power of the model in specific subgroups such as patients with grade 1 tumors or invasive lobular cancers for instance.

These findings support the use of RlapsRisk BC as a decision-support tool to refine treatment pathways. By identifying patients at low risk of recurrence, the model may help avoid unnecessary chemotherapy or overtreatment. Conversely, it can help flag patients at higher risk who may benefit from therapeutic escalation. This risk-based personalization of treatment aligns with evolving clinical practice trends, which emphasize risk stratification to inform treatment pathways.

It is also worth noting that multigene signatures such as Oncotype DX or EndoPredict are currently recommended primarily for patients with ambiguous clinicopathologic profiles—their widespread adoption is often limited by cost, and implementation complexity. RlapsRisk BC, in contrast, leverages digitized H&E-stained pathology slides and clinical data, offering a scalable and accessible alternative that could broaden risk stratification capacity across healthcare systems. Its added value over clinical variables, combined with lower barriers to access, supports broad applicability across the ER+/HER2– population.

Beyond prognostic accuracy, interpretability is key for clinical adoption. Prior analyses showed the AI model draws on morphological features linked to recurrence risk, such as nuclear atypia, mitotic activity, and stromal patterns [4]. Ongoing pathology-focused research aims to map predictive image regions to known histological features and identify novel tissue biomarkers, enhancing biological insight and trust in AI-driven pathology.

In summary, this blind validation study provides robust evidence that RlapsRisk BC delivers clinically actionable, and reproducible risk information in early-stage ER+/HER2– breast cancer. Its ability to refine patient stratification, outperform clinical models, and complement or substitute genomic assays, positions it as a valuable tool for guiding adjuvant treatment decisions. Further studies will be necessary to confirm its impact and optimize its integration into routine care. These results highlight the potential of AI-based pathology to support more personalized, data-driven treatment pathways for early ER+/HER2- breast cancer patients.

## Additional Information

## Acknowledgements

The authors thank the patients who participated and who did not oppose this additional research.

The authors also thank all the participating centers in the CANTO trial within the UNICANCER R&D frame: Gustave Roussy, Institut Jean Godinot, Institut de Cancérologie de Montpellier, Centre Oscar Lambret, Centre GF Leclerc, Institut de Cancérologie de Lorraine, Centre Léon Bérard, Centre François Baclesse, Centre Jean Perrin, Centre Antoine Lacassagne, Centre Paul Strauss, Institut Ste Catherine. The CANTO research project was supported by the French Government under the “Investment for the Future” program managed by the National Research Agency (ANR), grant no. ANR-10-COHO-0004.

Anonymised data were provided by the Wales Cancer Bank (DOI: http://doi.org/10.5334/ojb.46), which is supported by Health and Care Research Wales. It is possible that other researchers have also received materials from the same donors.

The TGCA-BRCA dataset cited in this study was generated by the TCGA Research Network: https://www.cancer.gov/tcga.

The authors gratefully acknowledge the Barts Cancer Institute Breast Biobank for their role in the collection and provision of tissue samples and associated data. We are especially thankful to the patients who generously donated their tissue and shared their data, enabling the completion of this work.

The authors gratefully acknowledge MD Anderson Cancer Centre for their role in the collection and provision of tissue samples and associated data.

## Authors’ contributions

M.L.-T., F.A., I.G., V.G., V.A and Z.V conceived the project. Data Curation and collection was conducted by I.G, M.L.T, E.H, S.K., B.V.D.V, L.J, C.M and A.V.S. Formal analysis was performed by V.G. and D.J. The original draft manuscript was written by V.G and all authors reviewed and edited the manuscript. V.G, Z.V, V.A, C.B, L.J., J.G, A.F., B.V.D.V, E.H, C.M., D.J, L.K.S., A.L.M., D.D., S.E., L.G, M.S, I.G, F.A, A.V.S, S.K and M.L.T. contributed to manuscript revisions and approved the final version. All authors read and approved the final manuscript.

## Ethics approval and consent to participate

This study complies with all relevant ethical regulations. All datasets comprised both consented patient data and data used under institutional ethics committee waivers.

## Consent for publication

NA.

## Data availability

The GrandTMA dataset that supports the findings of this study is available from Gustave Roussy but restrictions apply to the availability, which was used with permission for the current study, and so is not publicly available. The CANTO dataset used for training is available from UNICANCER but restrictions apply to the availability of data, which was used with permission for the current study, and so is not publicly available. The datasets, or a test subset, may be available from Gustave Roussy or UNICANCER, subject to ethical approvals. Requests for data access can be submitted to https://redcap.link/DataRequestClinicalTrialsGustaveRoussy for GrandTMA or directed to for CANTO cohorts.

The datasets used in this study, including GrandTMA, CANTO, CYPATH, CARDIFF, GRONINGEN, CURIE, BARTS, MDA, EP-GR, and OD-GR, are proprietary data of the respective medical centers and institutions. Access to these datasets, or subsets thereof, may be available upon request and subject to ethical approvals and data use agreements with the originating centers. Interested researchers should contact the individual institutions for specific data access procedures.

## Code availability

The code used for training the models has a large number of dependencies on internal tooling and its release is therefore not feasible. However, all experiments and implementation details are described thoroughly in the “Methods” and Supplementary Information (Cross-validation experiments and model hyperparameters) so that they can be independently replicated with non-proprietary libraries.

## Competing interests

V.G., Z.V., V.A., J.G, A.F, M.B, E.H, D.J, L.G, M.S are employees of Owkin Inc.

F.A. declares institutional financial interests, research grants with Novartis, Pfizer, AstraZeneca, Eli Lilly, Daiichi, Roche, and Sanofi. B.P. reports Consulting fees from Astra Zeneca (institutional), Seagen (institutional), Gilead (institutional), Novartis (institutional), Lilly (institutional), MSD (institutional), Pierre Fabre (personal), Daiichi-Sankyo (institutional/personal); research funding (institutional) from Astra Zeneca, Daiichi-Sankyo, Gilead, Seagen, MSD, and Fondation ARC. Travel support: Astra Zeneca; Pierre Fabre; MSD; Daiichi-Sankyo.

M.L.-T. reports consulting fees from Astra Zeneca (institutional/personal), Seagen (personal), Lilly (personal), MSD (institutional/personal), Pierre Fabre (personal), Daiichi-Sankyo (institutional/personal), Myriad Genetics (personal), Exact Sciences (personal), Roche Diagnostics ((institutional/personal); research funding (institutional) from Roche Diagnostics, Daiichi-Sankyo, and Pierre Fabre. Travel support: AstraZeneca, Seagen, and Daiichi-Sankyo. The remaining authors declare no competing interests.

All other co-authors declare no competing interests (D.D., C.B., L.J., B.V.D.V, C.M, L.K.S, A.-L. M., S.E, I.G, A.V-S, S.K)

## Funding information

RlapsRisk BC is being developed under the guidelines of PortrAIt, a french consortium financed by the government within the framework of France 2030 and by the European Union - Next Generation EU within the framework of the France Relance Plan.

## Supplementary material

**Supplementary Figure 1.**
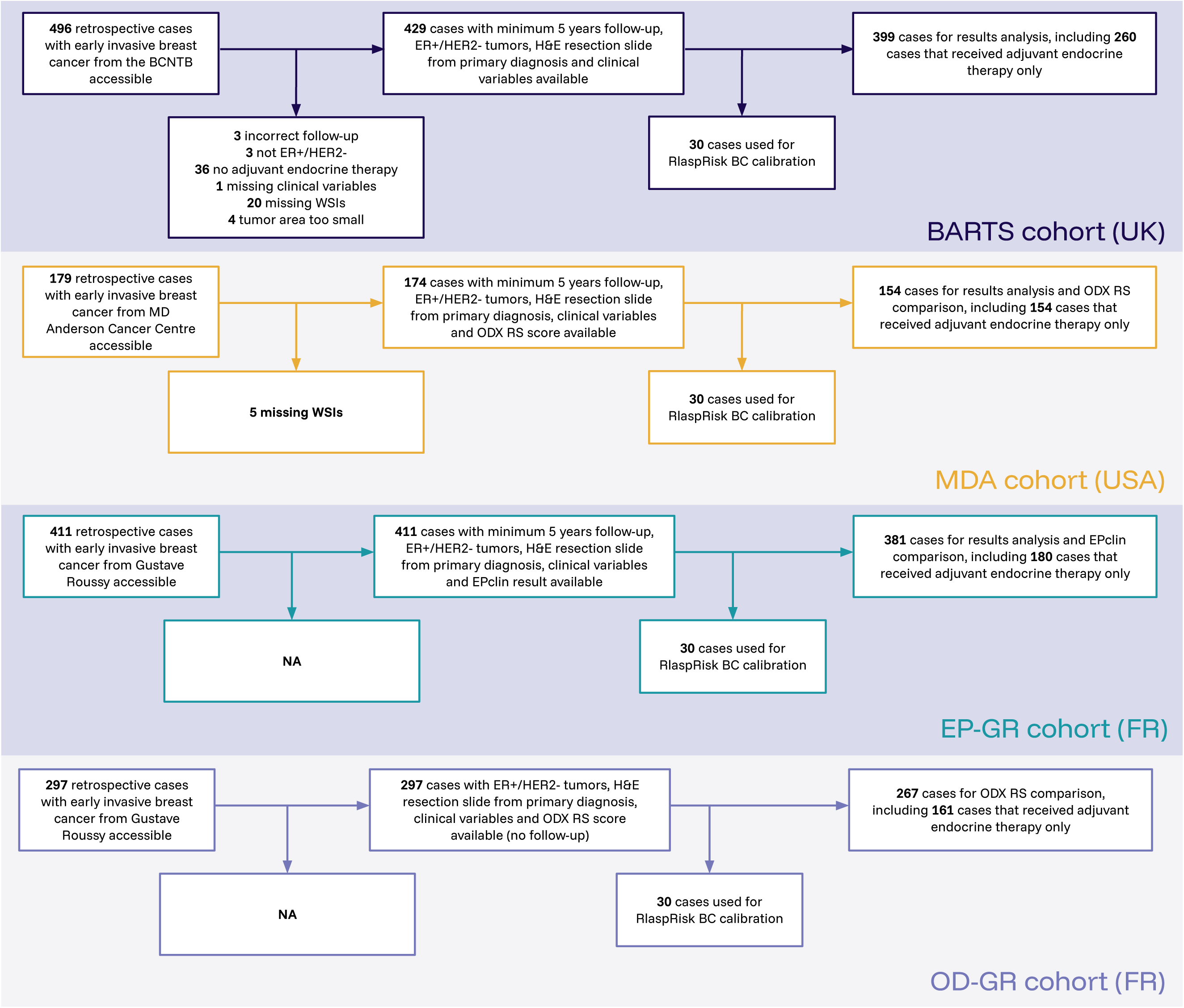

**Supplementary Figure 2.**
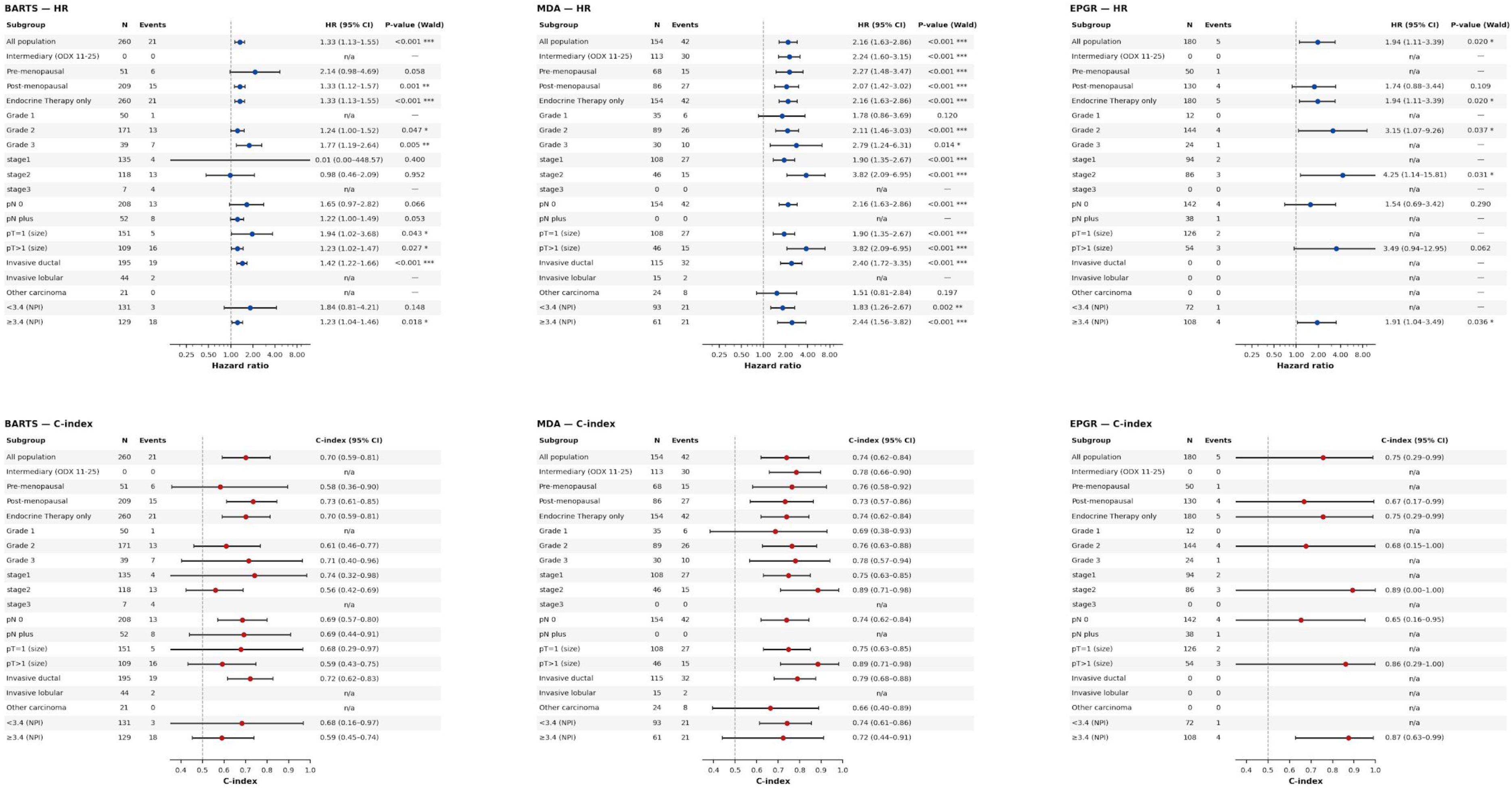

**Supplementary Figure 3.**
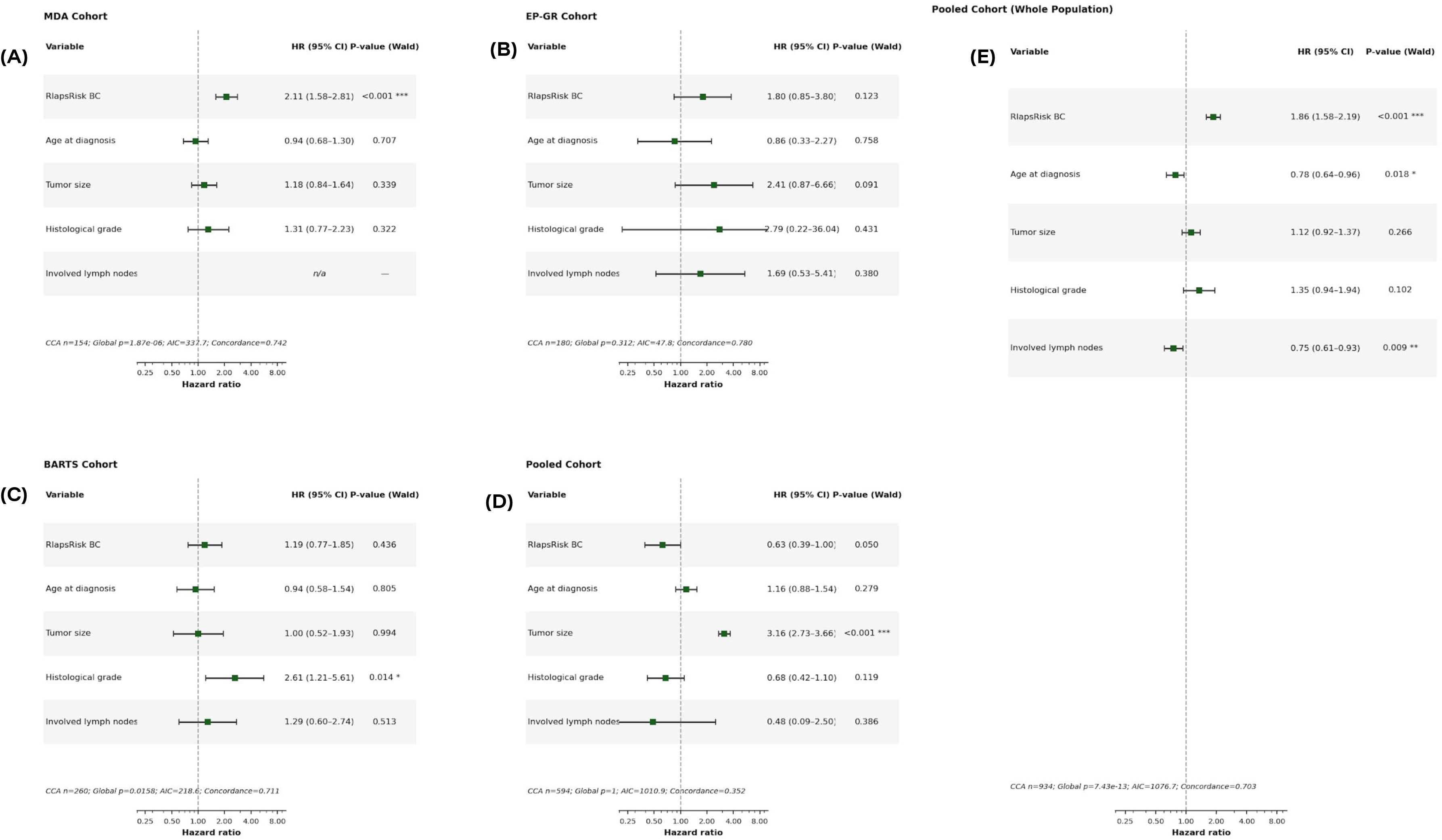

**Supplementary Figure 4.**
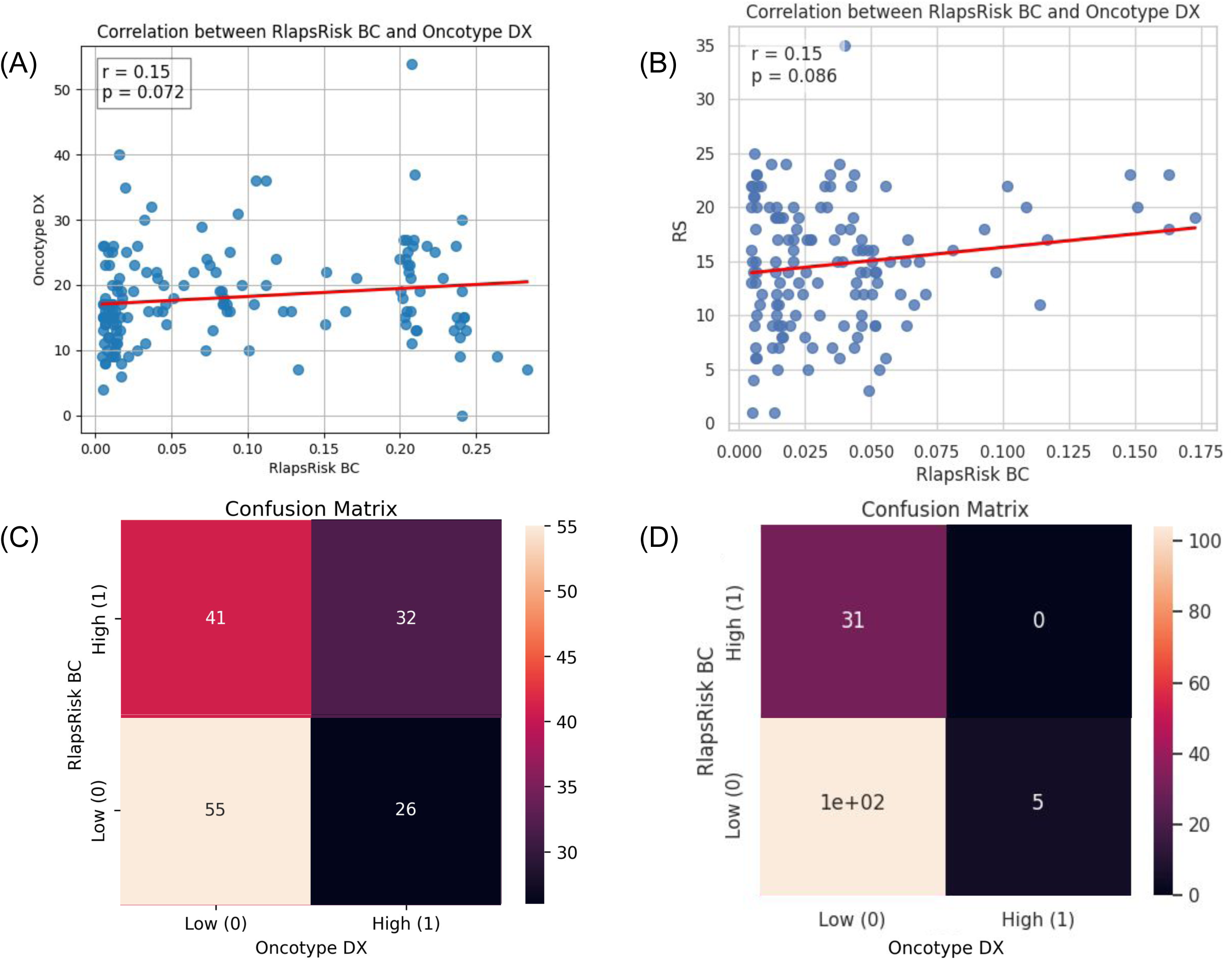

**Supplementary Figure 5.**
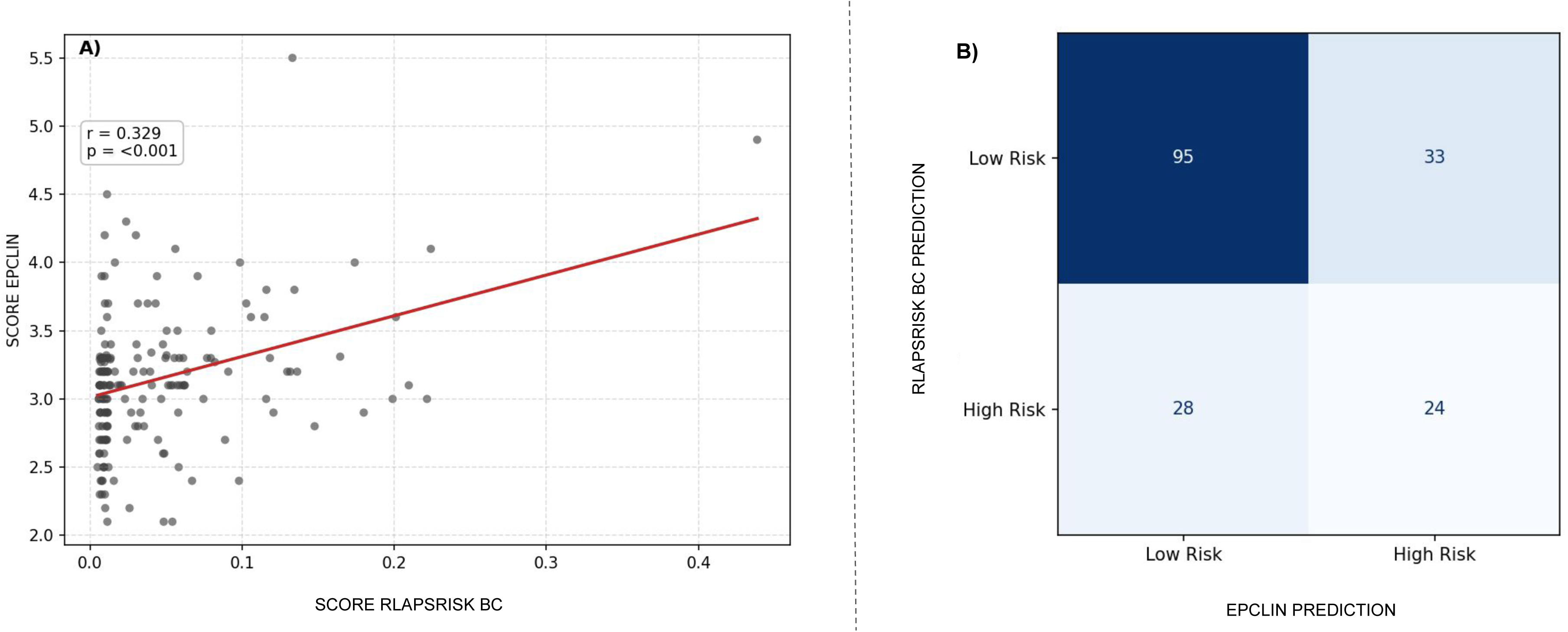

**Supplementary Figure 6.**
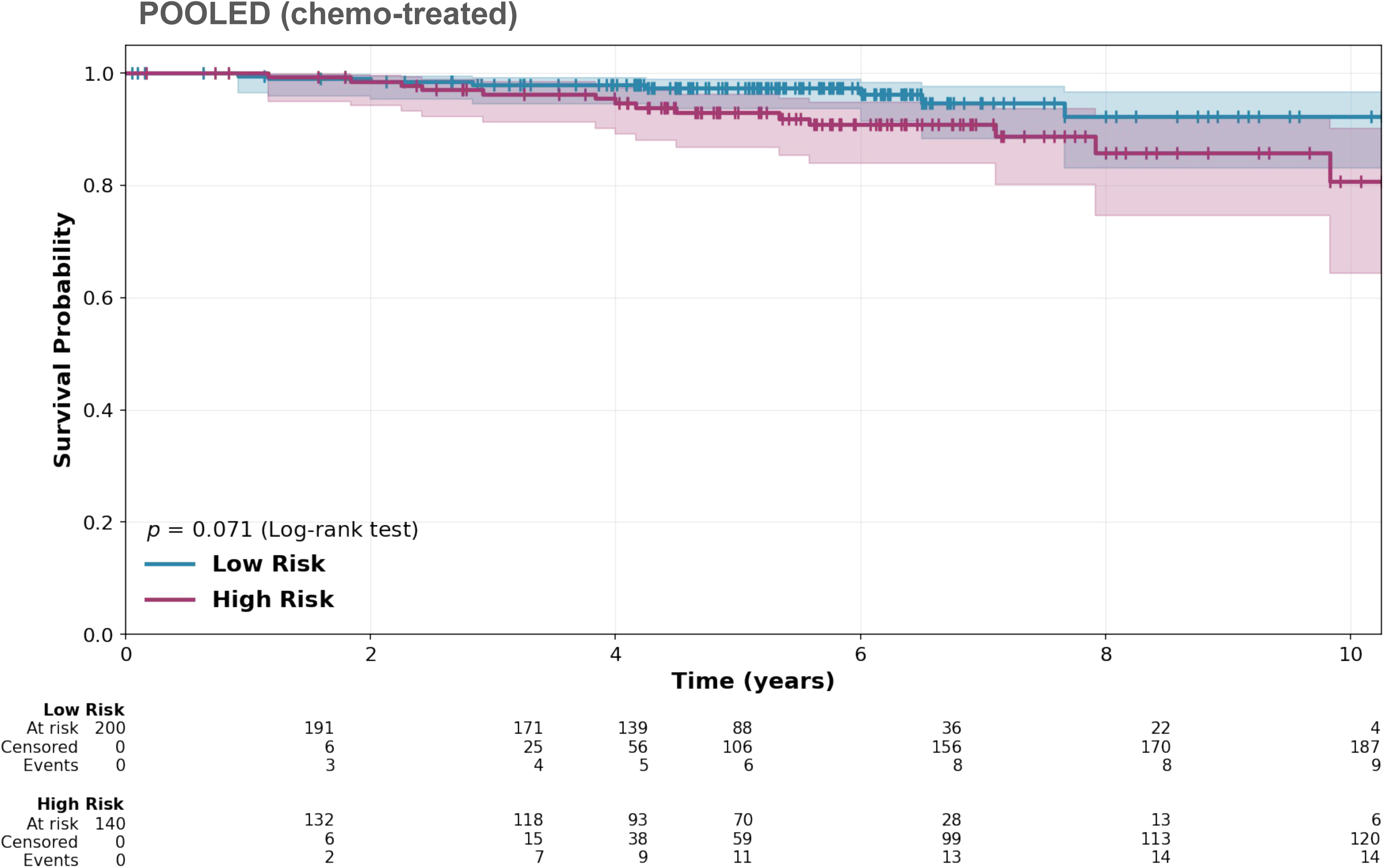

**Supplementary Figure 7.**
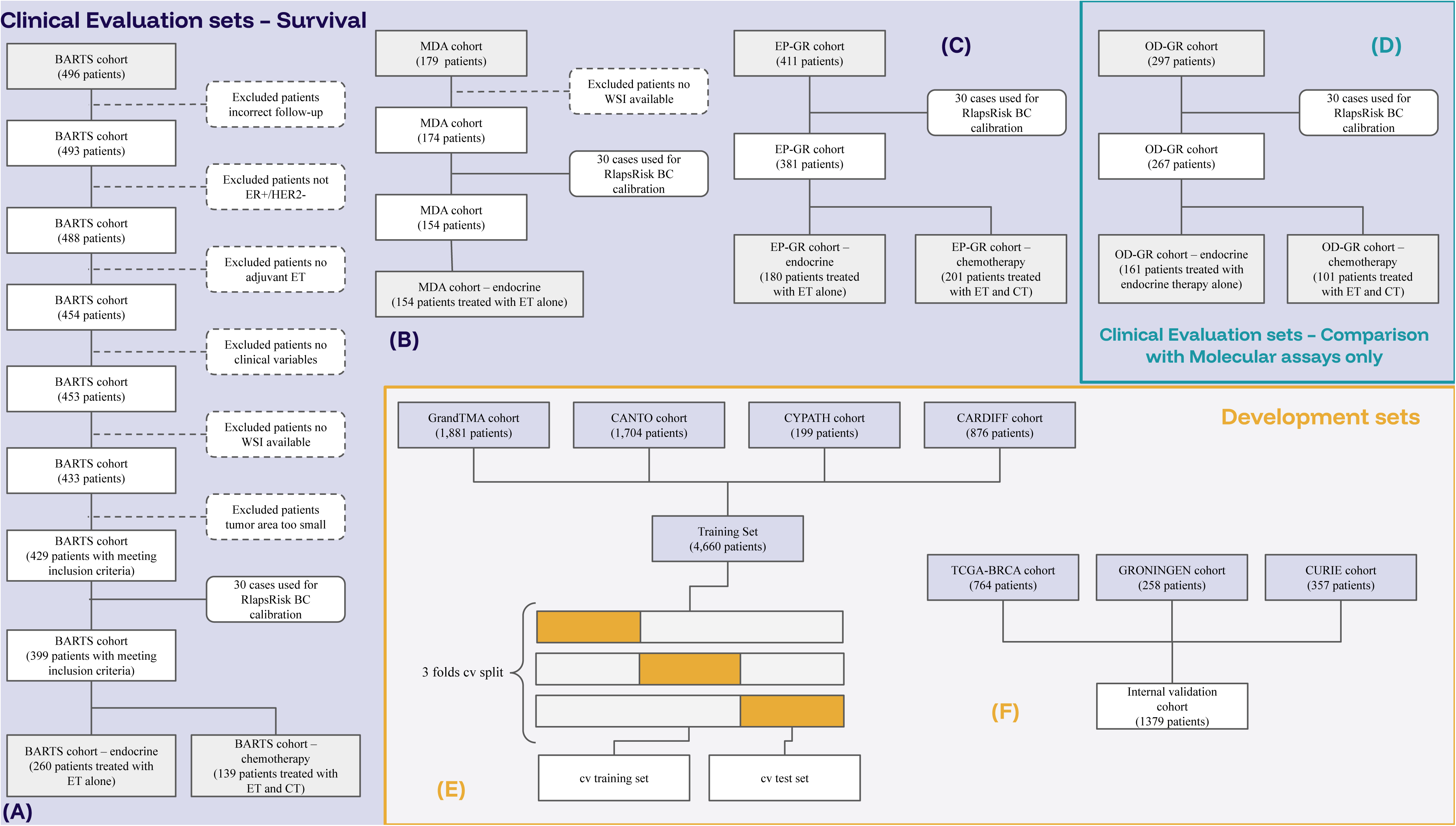

## Notes

### Funding Statement

This study did not receive any funding

### Author Declarations

In each country, the ethical approvals or waivers were obtained from the relevant institutional review boards or ethics committees for each participating study. These retrospective studies were conducted across sites with varying regulatory and ethical requirements. Informed consent was obtained from patients where required by local regulations; in some studies, consent was waived due to the retrospective nature of the data. The datasets used in our study were individual-level data and that all individual-level data had been de-identified prior to its use in this study.

### Summary of Updates

- Refinement of Comparative Terminology: We replaced the term "gold standard" with specific references to Oncotype DX and EndoPredict, while adding a discussion on the inherent inconsistencies between these genomic assays. - Methodological Clarification: We clarified the inclusion criteria for slide analysis, specifying the minimum tumor area requirement (150 tumor tiles/1.9 mm2) and providing a flowchart in Supplementary Figure 7 to account for excluded slides. - Update of Results and Subgroup Analyses: All analyses were re-run to ensure accuracy. Results are now presented specifically for the ER+/HER2- target population. We added p-values to hazard ratios (HR) and confidence intervals to provide greater statistical transparency. - Discussion of Limitations: We acknowledged the limitations regarding the sample size of our external validation cohorts, particularly in subgroup analyses, and explicitly noted the need for larger cohorts to confirm performance in specific subgroups (e.g., Grade 1 tumors, invasive lobular cancers). - Consistent Formatting and Figures: We harmonized figure legends and abbreviations throughout the manuscript, corrected axes to reflect years rather than months where relevant, and ensured all cited supplementary figures accurately align with the text. - Re-contextualization of Clinical Utility: We tempered claims regarding clinical utility by clarifying that while RlapsRisk BC provides distinct prognostic signals and improves granularity in intermediate-risk patients, it serves as a promising complement or alternative to existing genomic tests rather than a standalone replacement.

