## Supplementary Materials for "Clinical Validation of RlapsRisk BC in an international multi-cohorts setting"


**Supplementary Materials** **1**

Tables 2

Supplementary Table 1: Patients characteristics in the BARTS Cohort. Abbreviations: dRFI: distant recurrence-free interval. 3

Supplementary Table 2: Patients characteristics in the EP-GR Cohort. Abbreviations: dRFI: distant recurrence-free interval. 4

Supplementary Table 3: Comparison of Concordance Index (C-Index) and AUC Performance Between RlapsRisk BC and Established Clinical Models. 4

Supplementary Table 4: Prognostic Performance Comparison for RlapsRisk BC, Oncotype Dx (RS) and EndoPredict (EPclin) for cohorts where scores were available. 4

Supplementary Table 5: Prognostic Performance of RlapsRisk BC in external validation cohorts, Stratified by Treatment Type. 6

Figures 7

Supplementary Figure 1: Flowchart illustrating the inclusion and exclusion criteria applied to define the final study populations across the external validation cohorts. 7

Supplementary Figure 2: Subgroup Analyses of RlapsRisk BC Performance in Patients Treated with Endocrine Therapy Alone Across External Validation Cohorts. 8

Supplementary Figure 3: Multivariable Cox proportional hazards models assessing the independent prognostic value of multiple variables, including the RlapsRisk BC histological score, for distant recurrence-free interval (dRFI) across three external validation cohorts. 9

Supplementary Figure 4: Correlation and Concordance Between RlapsRisk BC and Oncotype DX (RS) Classifications in the MDA cohort (left panel) and OD-GR (right panel). 10

Supplementary Figure 5: Correlation and Concordance matrix Between RlapsRisk BC and EPclin Classifications in the EP-GR cohort. 10

Supplementary Figure 6: Kaplan-Meier curves for the dRFI endpoint in HR+/HER2- eBC treated only with adjuvant endocrine therapy and chemotherapy (no patients treated with adjuvant endocrine therapy only) in the POOLED cohort. 11

Supplementary Figure 7: CONSORT Diagram describing the complete datasets used in this study. This flowchart illustrates the patient selection process, dataset categorization, and analysis strategy used to develop, calibrate, and evaluate the RlapsRisk BC prognostic model for ER-positive, HER2-negative breast cancer patients. 12

Methods 13

Feature Extraction 13

Model architecture and training procedures 13

Calibration 14

Integration of SCPC variables in the Multi-variable model and stratification thresholds 14

### Tables

| BARTS cohort | | | | | |
|---|---|---|---|---|---|
| Variable | Characteristics | ALL TREATMENTS | | Adjuvant Endocrine Therapy Alone | |
| Number of patients | | 399 | 100.00% | 260 | 100.00% |
| Age (in years) | 18-49 | 95 | 23.81% | 38 | 14.62% |
| | 50-59 | 127 | 31.83% | 83 | 31.92% |
| | 60-69 | 113 | 28.32% | 85 | 32.69% |
| | >70 | 64 | 16.04% | 54 | 20.77% |
| | mean ± SD | 57.60 ± 12.02 | | 60.39 ± 11.34 | |
| | [min ; max] | [28.00 ; 86.00] | | [31.00 ; 86.00] | |
| | median (5th, 95th) | 57.00 (36.00, 78.10) | | 60.00 (41.95, 81.00) | |
| Histological Grade | Grade 1 | 53 | 13.28% | 50 | 19.23% |
| | Grade 2 | 245 | 61.40% | 171 | 65.77% |
| | Grade 3 | 101 | 25.31% | 39 | 15.00% |
| Nodal status | N- | 248 | 62.16% | 208 | 80.00% |
| | N+ | 151 | 37.84% | 52 | 20.00% |
| | pN1 | 118 | 29.57% | 46 | 17.69% |
| Tumor Size | pT1 | 160 | 40.10% | 139 | 53.46% |
| | pT2/3/4 | 239 | 59.90% | 121 | 46.54% |
| | mean ± SD | 27.46 ± 18.31 | | 22.49 ± 13.19 | |
| | [min ; max] | [6.00 ; 135.00] | | [6.00 ; 81.00] | |
| | median (5th, 95th) | 22.00 (10.00, 65.00) | | 18.00 (9.00, 50.00) | |
| dRFI events | Number of dRFI events | 42 | 10.53% | 21 | 8.08% |
| | Number of dRFI events at 5 years | 25 | 6.27% | 12 | 4.62% |
| | Number of dRFI events at 10 years | 41 | 10.28% | 21 | 8.08% |
| dRFI duration | mean ± SD | 91.86 ± 34.14 | | 90.48 ± 33.95 | |
| | [min ; max] | [2.00 ; 162.00] | | [2.00 ; 162.00] | |
| | median (5th, 95th) | 91.00 (31.70, 149.10) | | 88.00 (32.95, 148.05) | |

Supplementary Table 1: Patients characteristics in the BARTS Cohort. Abbreviations: dRFI: distant recurrence-free interval.

| EP-GR cohort | | | | | |
|---|---|---|---|---|---|
| Variable | Characteristics | ALL TREATMENTS | | Adjuvant Endocrine Therapy Alone | |
| Number of patients | | 381 | 100.00% | 180 | 100.00% |
| Age (in years) | 18-49 | 108 | 28.35% | 63 | 31.34% |
| | 50-59 | 104 | 27.30% | 62 | 30.85% |
| | 60-69 | 126 | 33.07% | 58 | 28.86% |
| | >70 | 43 | 11.29% | 18 | 8.96% |
| | mean ± SD | 57.21 ± 10.84 | | 55.48 ± 10.78 | |
| | [min ; max] | [24.93 ; 86.72] | | [24.93 ; 76.59] | |
| | median (5th, 95th) | 58.29 (40.10, 73.35) | | 56.00 (37.62, 71.73) | |
| Histological Grade | Grade 1 | 23 | 6.04% | 12 | 6.67% |
| | Grade 2 | 277 | 72.70% | 144 | 80.00% |
| | Grade 3 | 81 | 21.26% | 24 | 13.33% |
| Nodal status | N- | 283 | 74.28% | 142 | 78.89% |
| | N+ | 98 | 25.72% | 38 | 21.11% |
| | pN1 | 98 | 25.72% | 38 | 21.11% |
| Tumor Size | pT1 | 222 | 58.27% | 107 | 59.44% |
| | pT2/3/4 | 159 | 41.73% | 73 | 40.56% |
| | mean ± SD | 19.83 ± 9.41 | | 18.71 ± 7.47 | |
| | [min ; max] | [6.00 ; 90.00] | | [6.00 ; 50.00] | |
| | median (5th, 95th) | 18.00 (10.00, 35.00) | | 18.00 (10.00, 32.15) | |
| dRFI events | Number of dRFI events | 7 | 1.84% | 5 | 2.78% |
| | Number of dRFI events at 5 years | 5 | 1.31% | 4 | 2.22% |
| | Number of dRFI events at 10 years | 7 | 1.84% | 5 | 2.78% |
| dRFI duration | mean ± SD | 59.43 ± 17.61 | | 59.67 ± 17.59 | |
| | [min ; max] | [0.62 ; 87.67] | | [0.69 ; 87.67] | |
| | median (5th, 95th) | 61.84 (19.11, 82.03) | | 61.54 (16.82, 82.46) | |

Supplementary Table 2: Patients characteristics in the EP-GR Cohort. Abbreviations: dRFI: distant recurrence-free interval.

| POOLED COHORT (Endocrine Therapy Alone) | | | | |
|---|---|---|---|---|
| **Prognostic Metric** | **RlapsRisk BC** | **Clinical-only model** | **CTS5** | **PredictBreast** |
| c-index (95% CI) | 0.77 (0.71-0.84) | 0.58 (0.51-0.65) | 0.60 (0.53-0.68) | 0.25 (0.43-0.60) |
| AUC at 5 year s(95% CI) | 0.79 (0.69-0.87) | 0.55 (0.47-0.65) | 0.61 (0.54-0.71) | 0.56 (0.51-0.62) |

Supplementary Table 3: Comparison of Concordance Index (C-Index) and AUC Performance Between RlapsRisk BC and Established Clinical Models.

This table presents the prognostic discrimination (measured by the concordance index and the AUC) of RlapsRisk BC compared to other commonly used clinicopathologic-based models and tools, including the clinical-only model, CTS5, and Predict Breast. The comparison was performed on the POOLED validation cohort, and C-index and AUC values are reported with their corresponding 95% confidence intervals, where available.

| Endocrine Therapy Alone | | | | |
|---|---|---|---|---|
| **Metric** | **Cohort** | **RlapsRisk BC** | **RS** | **EPclin** |
| correlation | MDA | | 0.15 (p= 0.072) | n/a |
| | OD-GR | | 0.15 (p= 0.086) | n/a |
| | EP-GR | | n/a | 0.33 (p<0.001) |
| Hazard Ratios (95% CI, p-value) | MDA | 7.54 (3.34-16.99) p=<0.001 | 0.98 (0.53-1.83) p=0.959 | n/a |
| | EP-GR | 9.02 (0.99-82.09) p=0.051 | n/a | 2.87 (0.47-17.56) p=0.25 |
| concordance index (95% CI) | MDA | 0.76 (0.68–0.83) | 0.54 (0.45–0.63) | n/a |
| | EP-GR | 0.77 (0.33-0.99) | n/a | 0.59 ( 0.11-1) |

Supplementary Table 4: Prognostic Performance Comparison for RlapsRisk BC, Oncotype Dx (RS) and EndoPredict (EPclin) for cohorts where scores were available.

P-values for hazard ratios were computed with a Wald test.

| Key results (dRFI endpoint) | All patients | | | | Patients treated with adjuvant endocrine treatment alone | | | Patients treated with adjuvant endocrine AND chemotherapy | | |
|---|---|---|---|---|---|---|---|---|---|---|
| | **POOLED** | **MDA** | **BARTS** | **EP-GR** | **POOLED** | **BARTS** | **EP-GR** | **POOLED** | **BARTS** | **EP-GR** |
| nTOT | 934 | 154 | 399 | 381 | 594 | 260 | 180 | 340 | 139 | 201 |
| ndRFI(%) | 91 (9.74%) | 42 (27.27%) | 42 (10.53%) | 7 (1.84%) | 68 (11.45%) | 21 (8.08%) | 5 (2.78%) | 23 (6.76%) | 21 (15.11%) | 2 (1.00%) |
| HR high- vs low- risk (95% CI, p-value) | 4.92 (3.14-7.72) p=<0.001 | 7.54 (3.34-16.99) p=<0.001 | 2.72 (1.48-5.00) p=0.001 | 9.88 (1.16-84.28) p=0.036 | 7.44 (4.33-12.78) p=<0.001 | 3.93 (1.65-9.34) p=0.002 | 9.02 (0.99-82.09) p=0.051 | 2.13 (0.92-4.92) p=0.078 | 1.53 (0.64-3.63) p=0.336 | N/A |
| Difference significance between high and low risks at 5 years after surgery (p-value) | <0.001 | <0.001 | 0.004 | 0.053 | <0.001 | 0.010 | 0.079 | 0.075 | 0.307 | N/A |
| Difference significance between high and low risks at 10 years after surgery (p-value) | <0.001 | <0.001 | 0.006 | <0.001 | <0.001 | 0.032 | 0.001 | 0.081 | 0.176 | N/A |
| Proportion of dRFI events at 5 years after surgery in the low risk group (95% CI) | 2.50 (1.49-4.18) | 4.73 (1.55-13.98) | 3.81 (2.07-6.96) | 0.44 (0.06-3.06) | 2.41 (1.26-4.58) | 2.61 (1.09-6.16) | 0.85 (0.12-5.86) | 2.67 (1.12-6.32) | 7.01 (2.98-16.02) | 0.00 (0.00-0.00) |
| Proportion of dRFI events at 10 years after surgery in the low risk group (95% CI) | 7.83 (5.03-12.10) | 13.42 (6.50-26.57) | 8.93 (5.51-14.31) | 0.44 (0.06-3.06) | 7.94 (4.70-13.27) | 7.59 (3.88-14.59) | 0.85 (0.12-5.86) | 7.66 (3.38-16.89) | 12.60 (6.42-23.94) | 0.00 (0.00-0.00) |
| Proportion of dRFI events at 5 years after surgery in the high risk group (95% CI) | 13.07 (9.67-17.54) | 34.74 (23.63-49.12) | 11.73 (7.24-18.70) | 3.74 (1.41-9.77) | 17.80 (12.70-24.65) | 11.24 (5.52-22.16) | 6.26 (2.06-18.16) | 7.07 (3.73-13.16) | 12.20 (6.29-22.92) | 2.08 (0.30-13.88) |
| Proportion of dRFI events at 10 years after surgery in the high risk group (95% CI) | 35.92 (27.37-46.18) | 70.03 (55.67-83.21) | 22.52 (14.30-34.42) | 51.87 (17.41-93.89) | 47.24 (35.56-60.55) | 19.35 (11.12-32.45) | 53.13 (11.43-99.12) | 19.29 (9.88-35.69) | 23.05 (12.73-39.58) | 51.04 (9.74-99.31) |
| C-index at 5 years (95% CI) | 0.76 (95% CI, 0.69-0.83) | 0.78 (95% CI, 0.68-0.86) | 0.69 (95% CI, 0.57-0.80) | 0.76 (95% CI, 0.40-0.98) | 0.77 (95% CI, 0.68-0.85) | 0.69 (95% CI, 0.51-0.85) | 0.77 (95% CI, 0.25-0.99) | 0.71 (95% CI, 0.55-0.87) | 0.66 (95% CI, 0.47-0.84) | 0.69 (95% CI, 0.62-0.77) |
| C-index difference with SCPC model at 5 years | 0.17 (p-val < 0.001) | 0.19 (p-val=0.002) | 0.06 (p-val > 0.05) | 0.14 (p-val > 0.05) | 0.20 (p-val < 0.001) | 0.07 (p-val > 0.05) | 0.05 (p-val > 0.05) | -0.01 (p-val > 0.05) | 0.07 (p-val > 0.05) | 0.55 (p-val = 0) |
| C-index at 10 years (95% CI) | 0.75 (95% CI, 0.69-0.81) | 0.76 (95% CI, 0.67-0.83) | 0.70 (95% CI, 0.61-0.79) | 0.76 (95% CI, 0.41-0.98) | 0.77 (95% CI, 0.70-0.84) | 0.71 (95% CI, 0.57-0.83) | 0.76 (95% CI, 0.20-0.99) | 0.71 (95% CI, 0.57-0.84) | 0.66 (95% CI, 0.51-0.80) | 0.76 (95% CI, 0.63-1.00) |
| C-index difference with SCPC model at 10 years | 0.17 (p-val < 0.001) | 0.18 (p-val < | 0.04 (p-val > | 0.14 (p-val > | 0.19 (p-val = 0) | 0.03 (p-val > | 0.04 (p-val > | 0.01 (p-val > 0.05) | 0.07 (p-val > | 0.65 (p-val=0. |

| Key results (dRFI endpoint) | All patients | | | | Patients treated with adjuvant endocrine treatment alone | | | Patients treated with adjuvant endocrine AND chemotherapy | | |
|---|---|---|---|---|---|---|---|---|---|---|
| | POOLED | MDA | BARTS | EP-GR | POOLED | BARTS | EP-GR | POOLED | BARTS | EP-GR |
| | | 0.001) | 0.05) | 0.05) | | 0.05) | 0.05) | | 0.05) | 004) |
| AUC at 5 years (95% CI) | 0.77 (95% CI, 0.69-0.84) | 0.78 (95% CI, 0.66-0.87) | 0.70 (95% CI, 0.57-0.81) | 0.73 (95% CI, 0.36-0.97) | 0.79 (95% CI, 0.69-0.87) | 0.70 (95% CI, 0.52-0.86) | 0.75 (95% CI, 0.19-1.00) | 0.71 (95% CI, 0.54-0.86) | 0.67 (95% CI, 0.47-0.85) | 0.71 (95% CI, 0.62-0.79) |
| AUC difference with SCPC model at 5 years | 0.20 (p-val < 0.001) | 0.22 (p-val=0.005) | 0.06 (p-val > 0.05) | 0.17 (p-val > 0.05) | 0.23 (p-val < 0.001) | 0.08 (p-val > 0.05) | 0.04 (p-val > 0.05) | 0.02 (p-val > 0.05) | 0.06 (p-val > 0.05) | 0.56 (p-val = 0) |
| AUC at 10 years (95% CI) | 0.80 (95% CI, 0.72-0.86) | 0.79 (95% CI, 0.67-0.89) | 0.75 (95% CI, 0.65-0.84) | n/a | 0.82 (95% CI, 0.74-0.89) | 0.77 (95% CI, 0.64-0.89) | n/a | 0.74 (95% CI, 0.59-0.87) | 0.74 (95% CI, 0.59-0.88) | n/a |
| AUC difference with SCPC model at 10 years | 0.28 (p-val = 0) | 0.29 (p-val=0.001) | 0.09 (p-val > 0.05) | n/a | 0.27 (p-val < 0.001) | 0.03 (p-val > 0.05) | n/a | 0.25 (p-val=0.011) | 0.18 (p-val > 0.05) | n/a |
| Sensitivity at 5 years | 73.28% | 85.83% | 60.14% | 80.27% | 76.34% | 58.53% | 74.43% | 64.98% | 61.62% | 100.00% |
| Sensitivity at 10 years | 70.91% | 83.33% | 53.71% | n/a | 72.05% | 44.16% | n/a | 67.12% | 62.38% | n/a |
| Specificity at 5 years | 68.90% | 63.22% | 69.68% | 69.95% | 73.97% | 78.38% | 73.53% | 59.91% | 53.72% | 66.67% |
| Specificity at 10 years | 75.86% | 65.52% | 79.31% | n/a | 81.48% | 90.38% | n/a | 62.86% | 62.86% | n/a |
| PPV at 5 years | 13.32% | 34.30% | 11.96% | 3.91% | 18.25% | 11.85% | 6.39% | 7.07% | 12.26% | 2.17% |
| PPV at 10 years | 38.49% | 63.79% | 28.29% | n/a | 49.18% | 35.76% | n/a | 20.84% | 26.08% | n/a |
| NPV at 5 years | 97.53% | 95.22% | 96.23% | 99.57% | 97.62% | 97.44% | 99.16% | 97.33% | 93.02% | 100.00% |
| NPV at 10 years | 92.45% | 84.35% | 91.85% | n/a | 92.14% | 93.04% | n/a | 92.92% | 88.83% | n/a |

Supplementary Table 5: Prognostic Performance of RlapsRisk BC in external validation cohorts, Stratified by Treatment Type.

Metrics are based on distant recurrence-free interval (dRFI) events and include hazard ratio (HR), concordance index (C-index), AUC, sensitivity, specificity, positive predictive value (PPV), negative predictive value (NPV).
Note that MDA results are reported in the 'All patients' column only, as 100% of patients received adjuvant endocrine therapy alone.
P-values for comparisons of C-indexes and AUCs were computed with a Z-test.

### Figures

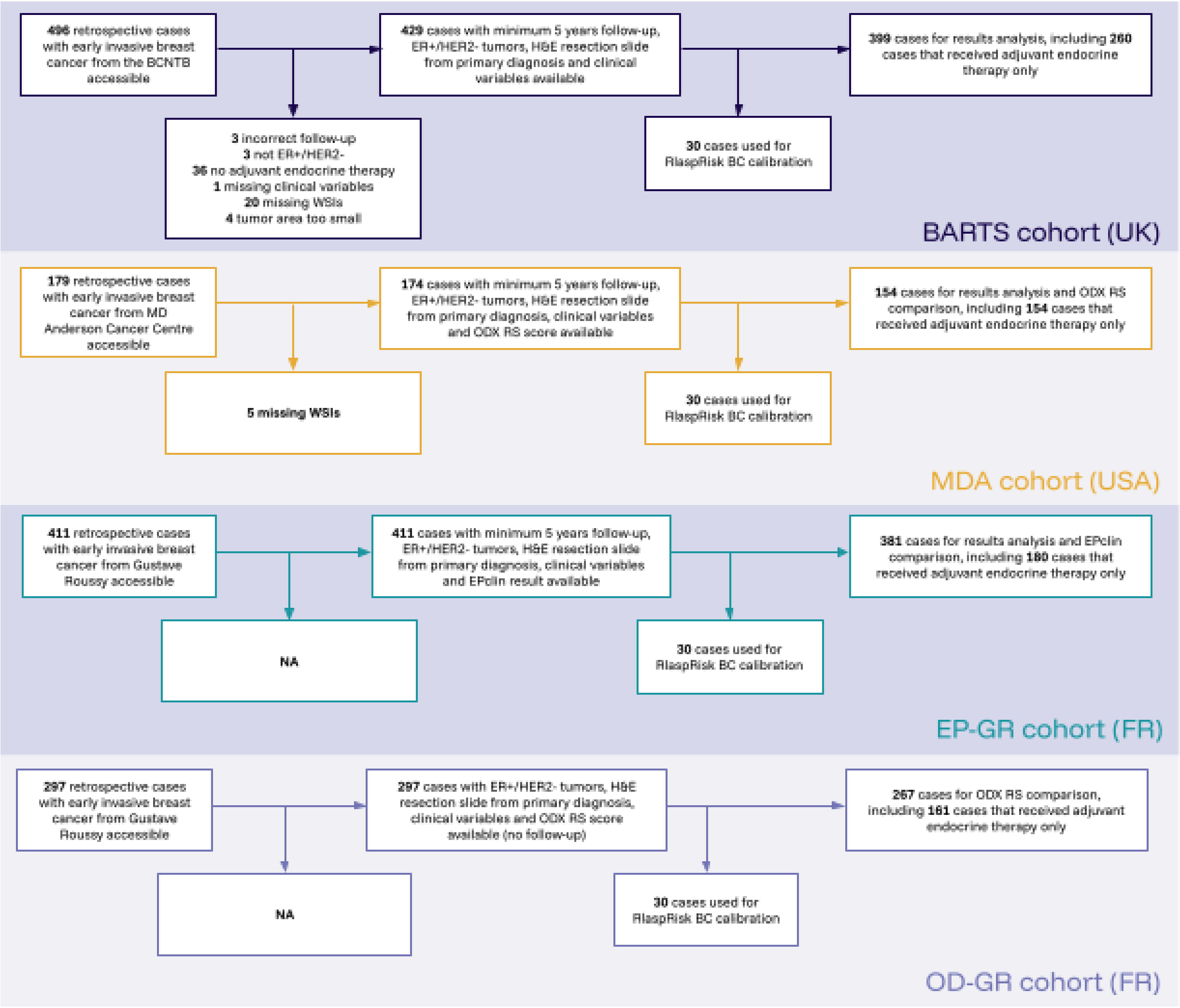


Supplementary Figure 1: Flowchart illustrating the inclusion and exclusion criteria applied to define the final study populations across the external validation cohorts.

The diagram details the sequential filtering steps used to construct the evaluation datasets for the BARTS, MDA, EP-GR, and OD-GR cohorts, including criteria such as availability of histological slides, complete clinical data, treatment information, and follow-up. This standardized selection process ensured consistency and comparability across cohorts for subsequent prognostic analyses.

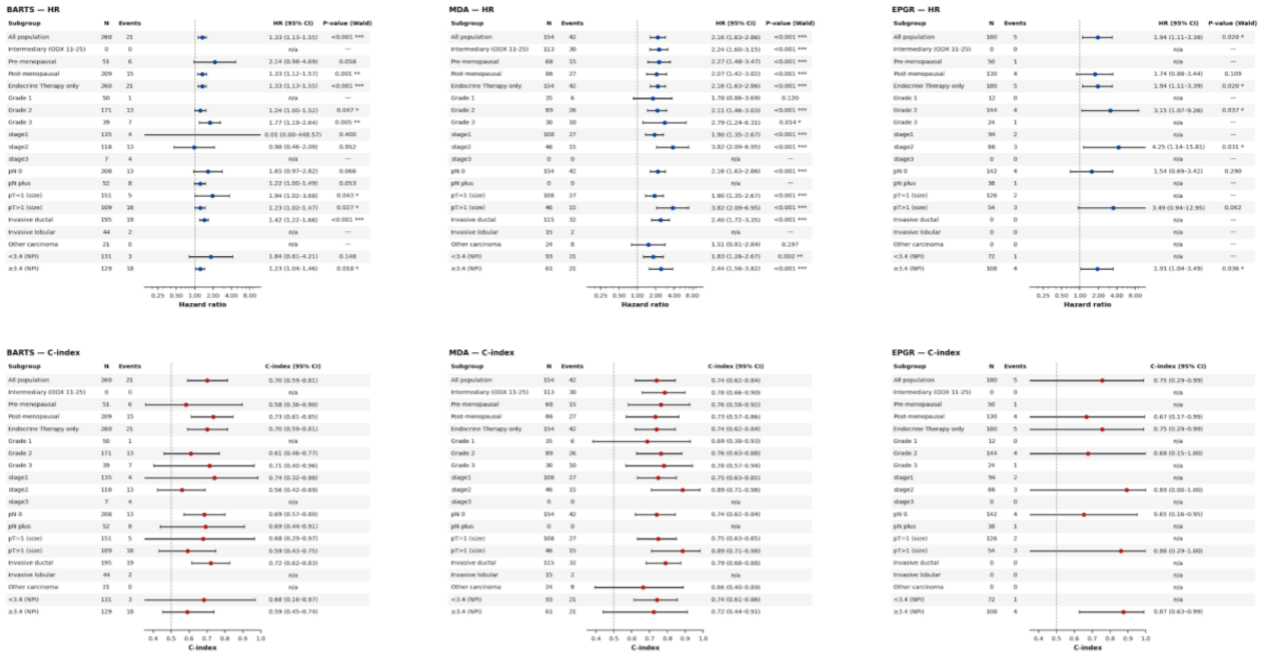


Supplementary Figure 2: Subgroup Analyses of RlapsRisk BC Performance in Patients Treated with Endocrine Therapy Alone Across External Validation Cohorts.

Forest plots display subgroup analyses for patients receiving endocrine therapy only in each validation cohort: BARTS (left panel), MDA (middle panel), and EP-GR (right panel).
The upper panels present hazard ratios (HRs) for distant recurrence-free interval (dRFI) comparing RlapsRisk BC high- vs low-risk groups within each clinical subgroup. The lower panels show the corresponding concordance indices (C-indices) with 95% confidence intervals, reflecting the discriminative performance of the score within each subgroup.
HR p-values were computed with a Wald test.

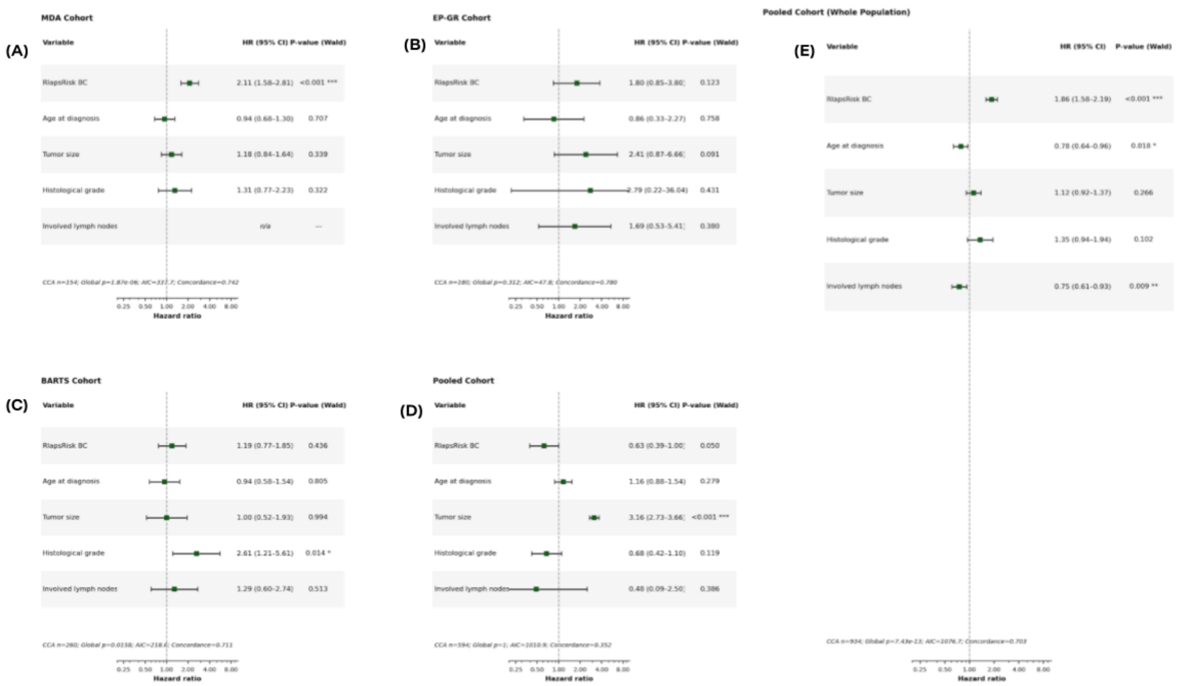


Supplementary Figure 3: Multivariable Cox proportional hazards models assessing the independent prognostic value of multiple variables, including the RlapsRisk BC histological score, for distant recurrence-free interval (dRFI) across three external validation cohorts.

Forest plots display hazard ratios (HRs) with 95% confidence intervals for each covariate included in the models. Results are stratified by treatment group: the left panels correspond to patients treated with endocrine therapy alone, while the right panel presents results for the overall population (including patients treated with or without adjuvant chemotherapy) in the POOLED cohort. Specifically, the endocrine therapy-only subgroup includes the MDA cohort (top left), EP-GR cohort (top right), BARTS cohort (bottom left), and POOLED cohort (bottom right). On the right, the full POOLED cohort is shown. These analyses consistently demonstrate the independent prognostic contribution of RlapsRisk BC when adjusted for conventional clinicopathological variables across all cohorts.

HR p-values were computed with a Wald test.

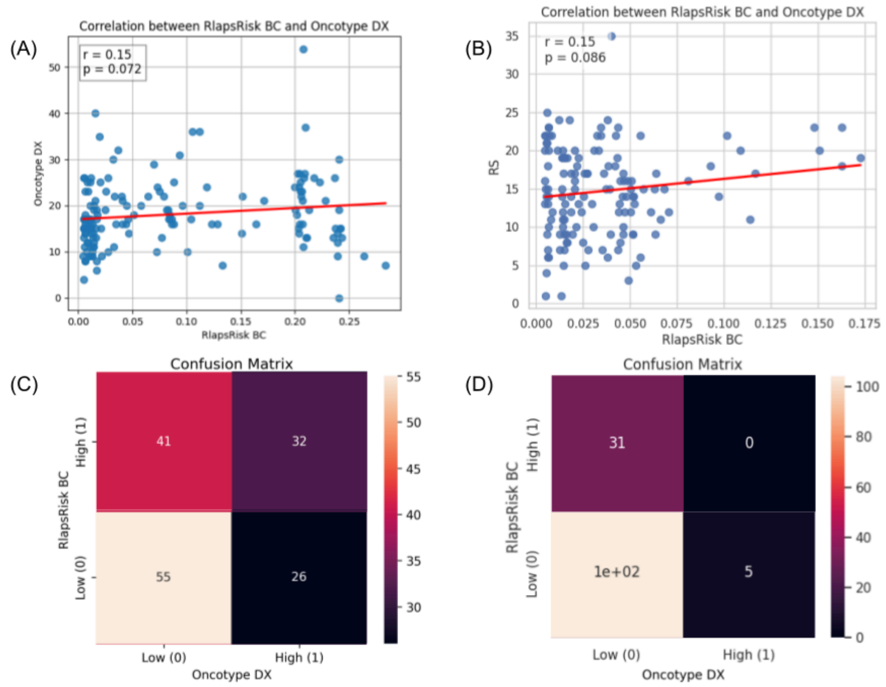


**Supplementary Figure 4**: Correlation and Concordance Between RlapsRisk BC and Oncotype DX (RS) Classifications in the MDA cohort (left panel) and OD-GR (right panel).

This figure presents scatter plots illustrating the correlation between RlapsRisk BC scores and Oncotype DX Recurrence Scores (RS), along with a confusion matrix summarizing the concordance in risk group classification (high vs low) between the two tools in the MDA cohort.
Correlation p-values were computed with a two-tailed t-test.

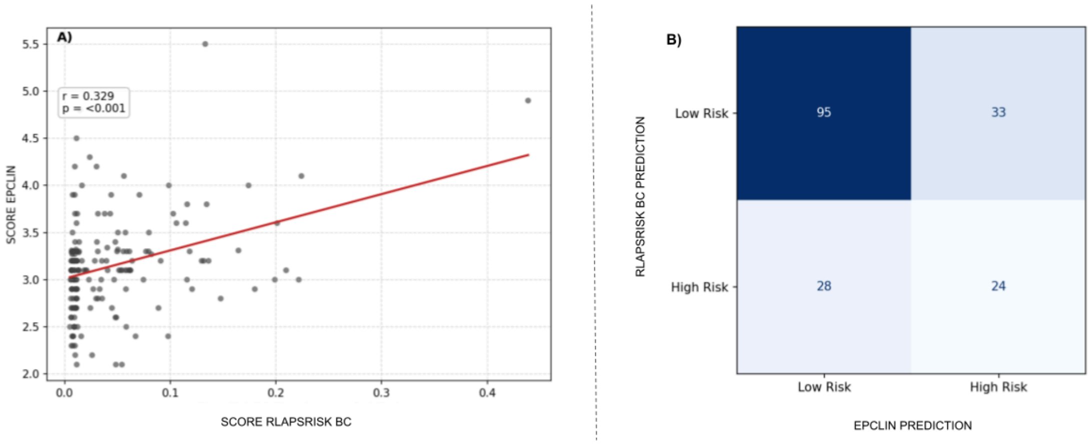


Supplementary Figure 5: Correlation and Concordance matrix Between RlapsRisk BC and EPclin Classifications in the EP-GR cohort.

This figure presents scatter plots illustrating the correlation between RlapsRisk BC scores and EPclin, along with a confusion matrix summarizing the concordance in risk group classification (high vs low) between the two tools in the EP-GR cohort.
Correlation p-value was computed with a two-tailed t-test.

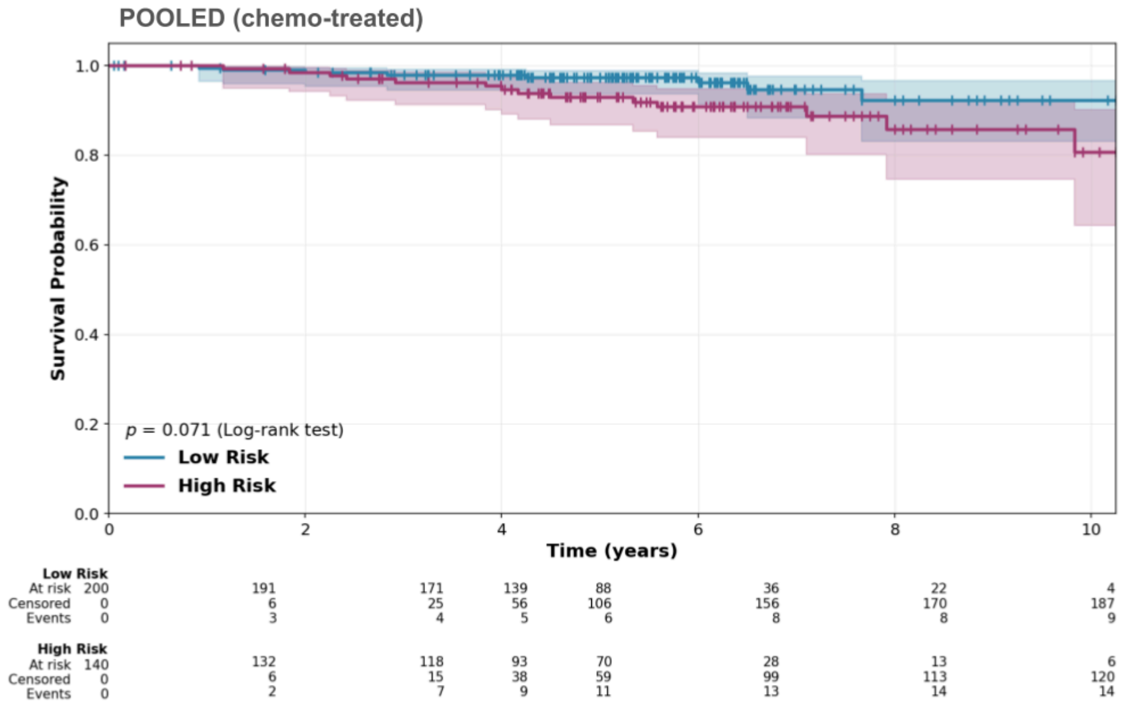


Supplementary Figure 6: Kaplan-Meier curves for the dRFI endpoint in HR+/HER2- eBC treated only with adjuvant endocrine therapy and chemotherapy (no patients treated with adjuvant endocrine therapy only) in the POOLED cohort.

Stratification p-value was computed with a logrank test.

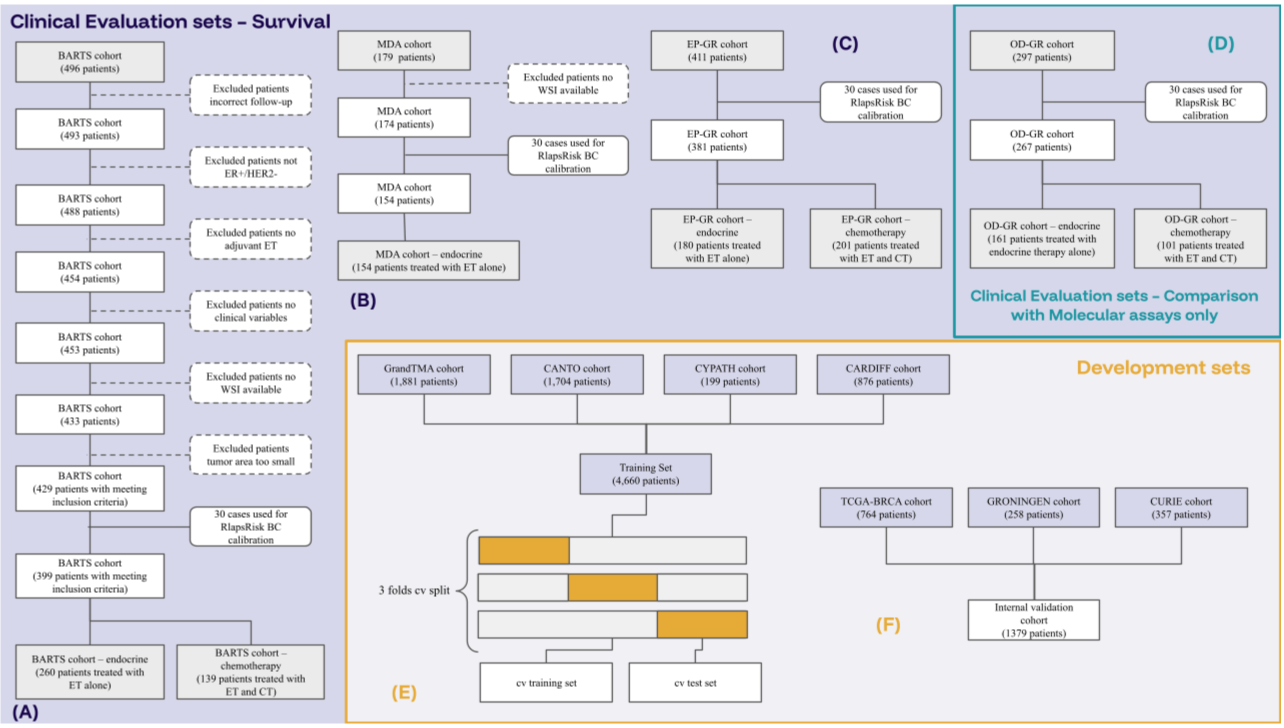


Supplementary Figure 7: CONSORT Diagram describing the complete datasets used in this study. This flowchart illustrates the patient selection process, dataset categorization, and analysis strategy used to develop, calibrate, and evaluate the RlapsRisk BC prognostic model for ER-positive, HER2-negative breast cancer patients.

Abbreviations: ET: endocrine therapy; CT: chemotherapy.

### Methods

#### Feature Extraction

The N tiles were embedded into D-dimensional feature vectors using a pre-trained Vision Transformer. We implemented iBOT ViT-B, a self-supervised learning transformer framework that improved performance for various prediction tasks in previous studies trained on the Cancer Genome Atlas PanCancer40M dataset, which covers 13 anatomic sites and 16 cancer subtypes for 5,558 patients, representing a total of 6,093 slides. Multiple data augmentations (random cropping, random flips, color jitter, random grayscale, Gaussian blur) were applied while the model was optimized for 153,000 iterations (approximately 50 h) on 32 NVIDIA Tesla V100 graphics processing units (GPU). The following hyperparameters were used to train the model: teacher temperature was set to 0.04 with an initial value of 0.04 and 30 warm-up epochs. AdamW optimizer50 was used and learning rate linearly ramped up during the first ten epochs to its base value scaled with the total batch size according to: 0.0005 × batch-size/25651. The final learning rate was set to 0.000002 through a cosine schedule. This frozen pre-trained algorithm was then used to extract features during training and inference.

#### Model architecture and training procedures

The N feature vectors (from the N tiles of a slide) were then aggregated using a MIL model trained to predict distant relapse at 5 years with a stratified threefold cross-validation approach, repeated five times. For each split, five models were also trained with random initialization of the weights. Given the limited number of events, stratification was performed based on event occurrence to ensure a minimum number of events in each fold. For inference on the external cohorts, predictions were generated by ensembling all 75 trained models through output averaging.

We reimplemented the attention-based model called Attention-Based MIL (ABMIL) proposed by Ilse et al. ABMIL is a deep MIL framework designed for weakly supervised classification. It aggregates instance-level (the WSI's tiles in our case) features using an attention-based pooling mechanism, which assigns learned importance weights to each instance within a bag. This allows the model to focus on the most relevant instances for prediction. The weighted instance representations are then combined into a single bag-level feature vector, which is processed by a fully connected layer to generate the final classification output. This approach enhances interpretability while effectively handling variability within each bag. A linear layer with L neurons (L = 256 here) was applied to the embedded features followed by a Gated Attention layer with L hidden neurons. A MLP with 128 neurons was then applied to the output. To speed-up training and fit all the data in memory, only a random subset of 8000 tiles per WSI was used, while all tiles of a slide are processed for inference. ABMIL was trained utilizing a loss function calculated with a smoothed variant of the concordance index, as suggested by Mayr and Schmid in this study. We used a smooth, differentiable loss function based on a sigmoid approximation of the standard C-index indicator function. This smoothing introduces a parameter σ that controls the transition sharpness, enabling gradient-based optimization. The loss function is integrated into a gradient boosting framework, iteratively minimizing the smoothed empirical risk to improve model discrimination. This approach ensures that the learned biomarker combinations are directly aligned with survival prediction performance. We also incorporated a multi-loss mechanism in order to increase the robustness of the predictions with respect to stainings or scanner variations. When, multiple slides per patients were available, with introduced a penalization term for the variance of the predicted score across the different WSIs of the patients. The model was trained with the following hyperparameters: batch size = 128, learning rate = 0.001, and MLP dropout = 0.4 (a comprehensive list of hyperparameters is provided in the Supplementary Methods). Given the 7.69% relapse rate in our training cohort, this resulted in an average of 9.84 events per minibatch, providing adequate supervision for model optimization. While minibatches without observed events were possible, their occurrence was rare (~0.0036% probability). Although such batches do not directly contribute to differentiating survival outcomes, their impact was mitigated by the smoothed C-index loss, which allows gradient updates even when minibatches contain few events.

The models were trained for 15 epochs with a batch size of 128. Each epoch required approximately 26 s to complete, resulting in a total training time of 6 min and 30 s per fold. The entire training of the models with a five-times-repeated threefold cross-validation process with multiple random weight initializations took approximately 1 h 38 min. Training was conducted on a Tesla T4 Nvidia GPU using the PyTorch framework. The CPU memory usage for loading all features of the training data was 45GB, with an additional 13GB of GPU memory utilized during training.

#### Calibration

To guarantee the robustness of our stratification in high or low risk across diverse data acquisition protocols of the external validation cohorts (scanners, training methodologies), we implemented Uniform Piecewise Approximation (UPA) as a calibration strategy. Inspired by image processing's histogram matching, UPA aligns the unseen dataset's model prediction distribution with a predefined reference distribution.

The reference distribution was defined as the set of scores obtained from inference of RlapsRisk BC on the H&E samples of the training cohort, from which we derived the cumulative distribution.

For each new dataset or new laboratory setting, a set of 30 samples is selected, comprising 10, 15, and 5 samples, respectively, from histological grades 1, 2, and 3. This selection aims to mirror the histological grade distribution observed in the discovery dataset. Histological grade was considered as the main prognostic factor that is routinely captured by the pathologist's analysis of the tumor slide only. Thus, we ensure a good representativity of morphologies and an ease of implementation in clinical practice. These 30 samples are used to generate a new set of RlapsRisk BC predictions, from which we derived an estimated cumulative distribution of the unseen dataset (corresponding to the data acquisition protocol of the laboratory).

Next, a linear function is fitted to map the unseen set's cumulative distribution onto the reference distribution, essentially aligning the prediction "shapes," without impacting the relative order of patients' risk. This mapping function was then applied to calibrate any new prediction from the unseen set, ensuring consistency with the desired reference distribution and enhancing generalizability across acquisition protocols. By leveraging UPA, we could achieve consistent model predictions even when applied to data acquired differently from the reference set.

#### Integration of SCPC variables in the Multi-variable model and stratification thresholds

To develop the RlapsRisk BC multimodal prognostic score for predicting 5-year distant relapse and to obtain predicted probabilities of occurrence of distant events before 5 and 10 years, we fitted a Weibull AFT (Accelerated Failure Time) model on the training dataset, incorporating the following clinical covariates: patient age, tumor size, number of invaded lymph nodes, and the histological risk score described in the previous section. To avoid information leakage, the histological risk score was derived exclusively from the averaged predictions of the validation splits of the histological model ensembles. The conversion from a continuous risk score to a multimodal based estimated probability of relapse was used to identify the threshold of RlapsRisk BC corresponding to a probability of 5-year DRFI event of 5% defined by the Weibull Models. This 5% distant event rate threshold corresponds to the 5-year interpolation of an exponential model from the 10-year distant event of 10%, which is the most common output of the molecular signatures currently used in clinical practice. The identified threshold is fixed for any application of RlapsRisk BC.
